# Clinical impact and cost-effectiveness of the updated COVID-19 mRNA Autumn 2023 vaccines in Germany

**DOI:** 10.1101/2023.10.09.23296505

**Authors:** K Joshi, S Scholz, M Maschio, M Kohli, A Lee, K Fust, B Ultsch, N van de Velde, E Beck

**Author notes:** Corresponding author: Keya Joshi.

## Abstract

**Objectives:** To assess the potential clinical impact and cost-effectiveness of coronavirus disease 2019 (COVID-19) mRNA vaccines updated for Autumn 2023 in adults aged ≥60 years and high-risk persons aged 30-59 years in Germany over a 1-year analytic time horizon (September 2023--August 2024).

**Methods:** A compartmental Susceptible-Exposed-Infected-Recovered model was updated and adapted to the German market. Numbers of symptomatic infections, number of COVID-19 related hospitalisations and deaths, costs, and quality-adjusted life-years (QALYs) gained were calculated using a decision tree model. The incremental cost-effectiveness ratio of an Autumn 2023 Moderna updated COVID-19 (mRNA-1273.815) vaccine was compared to no additional vaccination. Potential differences between the mRNA-1273.815 and the Autumn Pfizer-BioNTech updated COVID-19 (XBB.1.5 BNT162b2) vaccines, as well as societal return on investment for the mRNA-1273.815 vaccine relative to no vaccination, were also examined.

**Results:** Compared to no Autumn vaccination, the mRNA-1273.815 campaign is predicted to prevent approximately 1,697,900 symptomatic infections, 85,400 hospitalisations, and 4,100 deaths. Compared to an XBB.1.5 BNT162b2 campaign, the mRNA-1273.815 campaign is also predicted to prevent approximately 90,100 symptomatic infections, 3,500 hospitalisations, and 160 deaths. Across both analyses we found the mRNA-1273.815 campaign to be dominant.

**Conclusions:** The mRNA-1273.815 vaccine can be considered cost-effective relative to the XBB.1.5 BNT162b2 vaccine and highly likely to provide more benefits and save costs compared to no vaccine in Germany, and to offer high societal return on investment.

## INTRODUCTION

Following 12 months of a decreasing incidence of infections with Severe Acute Respiratory Syndrome coronavirus 2 (SARS-CoV-2), in May 2023 the World Health Organization (WHO) ended the coronavirus disease 2019 (COVID-19) global health emergency [1]. Since the start of the COVID-19 pandemic, the epidemiological situation in Germany has transitioned from a pandemic to an endemic circulation of SARS-CoV-2. The dominant Omicron variants and the high levels of immunity in the population due to vaccinations and previous infections have led to significantly fewer cases of severe illness and long-term consequences than at the beginning of the pandemic. Nevertheless, as of 21 September 2023, there were more than 175,000 deaths in Germany attributed to COVID-19 [2]

With the shift from the pandemic to the endemic phase, the German Standing Committee on Vaccination (Ständige Impfkommission; STIKO) at the Robert Koch Institute (RKI) updated its COVID-19 vaccination guidelines. The guidelines include a primary immunisation for people aged 18 years and older and annual, variant-adapted booster vaccinations, preferably in autumn, for persons 60 years or older, persons with high-risk conditions, residents in long-term care facilities, immunocompromised patients and their relatives as well as medical personnel [3].

Since the beginning of vaccination in the winter of 2020-21, the primary goal of the STIKO COVID-19 vaccination recommendation has been the prevention of severe outcomes (COVID-19-related hospitalisations and deaths), the protection of medical and nursing staff as well as those with other highly exposed occupations against infections with SARS-CoV-2, the prevention of transmission and protection in settings with high proportion of high-risk persons and high risk of outbreaks, and the maintenance of state functions and public life [4]. Since 2023, the vaccination goals are protection from severe disease and the avoidance of long-term consequences of COVID-19 [5]. Since April 2023, entitlement to COVID-19 vaccinations for people insured with the statutory health insurance in Germany is subject to the provisions of the vaccination guideline passed by the Joint Federal Committee of Physicians and Health Insurance Funds (G-BA) based on the STIKO recommendation.

As SARS-CoV-2 evolved from the ancestral strain (Wuhan-Hu-1) into the Omicron variants that have circulated since January 2022, the COVID-19 vaccines were modified during the pandemic, and new bivalent COVID-19 mRNA vaccines containing antigens to both the ancestral strain and the Omicron BA.4/BA.5 sub-variants were developed. However, by the end of January 2023, XBB sub-variants had begun to dominate globally, and initial studies reported that the bivalent COVID-19 vaccines had low effectiveness against these sub-variants [6,7]. Therefore, in May 2023, the WHO Technical Advisory Group on COVID-19 Vaccine Composition recommended that COVID-19 vaccines be updated once more to monovalent versions with an XBB subvariant [8].

Since September 2023 two mRNA vaccines, encoding the viral spike protein of SARS-CoV-2 Omicron XBB.1.5, have been licensed in the European Union for active immunisation to prevent COVID-19 caused by SARS-CoV-2 in individuals 12 years of age and older [9] and are recommended by the STIKO in Germany [3]. The safety profile of the Moderna updated Spikevax XBB.1.5 vaccine (mRNA-1273.815) is consistent with previously authorised vaccines and is anticipated to be effective against current SARS-CoV-2 variants [10]. Each vaccine dose contains the same amount of total mRNA as the previous bivalent version of the vaccine [11], although the formulation of these two updated mRNA COVID-19 vaccines differs in several areas, such as dosage, lipid nano particles and presentation: the mRNA-1273.815 vaccine contains andusomeran, an mRNA molecule with instructions for producing a protein from the Omicron XBB.1.5 subvariant of SARS-CoV-2 [12], whereas the Pfizer-BioNTech updated Comirnaty XBB.1.5 vaccine (XBB.1.5 BNT162b2) contains raxtozinameran, an mRNA molecule that also produces a protein from the Omicron XBB.1.5 subvariant of SARS-CoV-2 [13].

The safety and efficacy of both mRNA-1273 and BNT162b2 in reducing the risk of SARS-CoV-2 infection in the general population and in adolescents have been demonstrated in numerous phase 3 clinical trials [14-16]. Subsequent observational studies have provided evidence of the protection offered by two doses of both mRNA vaccines against SARS-CoV-2 infection and against COVID-19 hospitalisation and hospital death [17-22], despite the emergence of variants [23-25], although several observational studies have demonstrated that the differences in formulation impact vaccine effectiveness [17-22].

Due to the profound economic burden of COVID-19, several studies reporting the treatment costs of COVID-19 and its impact on healthcare budgets in different regions as well as the global economy have recently been published [26]. However, the availability of two mRNA vaccines has proved to be a critical tool against COVID-19, and there are several studies of the cost-effectiveness of vaccination both in the United States [27] as well as in low- and middle-income countries. A systematic literature review showed that vaccination programs across the world would be cost-effective and even cost-saving compared to no vaccination at all, even when the efficacy of vaccines varied, and when only a specific age range was targeted [28].

Nevertheless, in view of the changes to the epidemiological, clinical, and financial dimensions of COVID-19 vaccines, it remains important to determine if the use of COVID-19 vaccines is cost-effective [29], to better inform policy-makers about the overall burden of COVID-19 and the efficient vaccination strategies.

### Study objectives

The objective of this analysis was to model the potential clinical impact and cost-effectiveness of the Moderna Autumn 2023 mRNA-1273.815 vaccine campaign in a target population of persons aged 60 years and older, and high-risk persons aged 30-59 years in Germany over 1 year from September 2023 to August 2024. Outcomes of the mRNA-1273.815 vaccine campaign were compared to no COVID-19 vaccination. In addition, the clinical impact and cost-effectiveness of an Autumn mRNA-1273.815 vaccination campaign were compared with a Pfizer-BioNTech Autumn XBB.1.5 BNT162b2 campaign.

## METHODS

### Overview

A previously developed [29] Susceptible-Exposed-Infected-Recovered (SEIR) model was adapted to assess the clinical impact of an mRNA-1273.815 vaccination campaign in Germany. A decision tree was then used to calculate, based on the predicted number of infections from the SEIR model, the numbers of symptomatic infections, COVID-19 related outpatient visits, COVID-19-related hospitalizations, COVID-19-related deaths, quality-adjusted life-years (QALYs) gained, as well as corresponding medical and societal costs for each vaccination strategy. Both parts of the model are described in more detail below. For each vaccination strategy, we estimated the clinical and economic impact, and cost-effectiveness via the incremental cost-effectiveness ratio (ICER), economically justifiable prices (EJP) assuming different willingness-to-pay (WTP) thresholds, as well as resulting return on investment (ROI) and benefit-cost-ratios (BCR) of each vaccination strategy. Using this model, we conducted two sets of analyses in which the estimated clinical outcomes and cost-effectiveness of an mRNA-1273.815 vaccine were compared with (1) no COVID-19 vaccination and (2) an XBB.1.5 BNT162b2 vaccine campaign.

### SEIR Model

We used an age-stratified SEIR compartmental model adapted from Shiri et al., which has been described elsewhere [29,30]. Briefly, we assumed individuals are initially susceptible (S) and move to an exposed or latent state (E) after an effective contact with an infectious individual. This rate of movement is dictated by an age-specific contact matrix [31] that incorporates masking behavior, reduction in social mobility in the early phase of the pandemic, and transmissibility of the virus. After the latent period has elapsed, exposed individuals become infectious (I). Individuals then move to a recovered state (R) once the infectious period has passed. We allow for waning of natural immunity, permitting people to move back to the susceptible compartment, and assume the rate of waning is the same regardless of vaccination status.

All individuals start the simulation in the unvaccinated S state from 31 January 2020, and move through the vaccinated states, primary series (e.g., completion of two doses), booster 1 (e.g., dose 3), and booster 2 (e.g., dose 4), and booster 3 (e.g., dose 5), according to uptake rates as vaccination coverage increases (see section 1.3.1 in the Supplemental Appendix), and booster doses become available. Individuals were permitted to receive a booster vaccine only after completing the primary series. The mRNA-1273.815 campaign was modelled for the 4-month period from September 2023 to December 2023 for individuals who received the primary series, irrespective of the quantity or type of boosters received previously [3]. Full model details including equations and inputs are described in the Technical Appendix of this publication and in a separate Technical Appendix developed by Kohli et al. [29].

### Population

For each analysis, we were interested in understanding the impact of vaccination in adults in Germany aged 60 years and older, as well as those aged 30-59 years with at-risk conditions, mirroring STIKO recommendations. A previous study estimated the approximate size of the population at greater risk of severe COVID-19 outcomes following infection [32]. Using this study, we estimated 21% of the population in Germany aged 30-59 years had at least one chronic condition that increased the risk of a severe outcome following COVID-19 infection.

### Vaccine coverage

To model the impact of different circulating variants on vaccine effectiveness (VE), we divided the model simulation into three periods: pre-Omicron (31 January 2020--21 November 2021); Omicron BA.1/2 (22 November 2021--9 May 2022); and Omicron BA.4/5 (after 9 May 2022). The dates for variant predominance for Germany were selected using data from CoVariants.org [33].

Vaccination coverage information was obtained from the RKI, the national public health institute in Germany, which provides estimates of (1) age stratified daily vaccination counts by vaccine type (i.e., primary series, booster 1) and (2) daily vaccination counts by manufacturer by vaccine type. These data were used to calculate the age-specific prior uptake for the primary series, booster 1, booster 2, and booster 3 for Germany. We assumed that no additional primary series or first boosters were delivered after 27 February 2022, and no additional second or third boosters were delivered after 21 December 2022. For the mRNA-1273.815 as well as XBB1.5 BNT162b2 campaign, we assumed coverage [34] and uptake [35] for at-risk individuals aged 30-59 years and adults aged 60 years and older to be the same as for the 2021/2022 influenza vaccine season in Germany.

### Residual vaccine efficacy

To estimate residual VE for the population at the start of the 1-year analytic time horizon beginning September 2023, we used the initial VE and monthly waning against infection and severe disease for previously administered vaccines, accounting for the vaccine type, dose (e.g., primary series, booster 1), and circulating variant at the time of administration.

### Autumn 2023 vaccine efficacy/effectiveness

Both the mRNA-1273.815 and XBB1.5 BNT162b2 Autumn 2023 vaccines were assumed to be well-matched to the circulating variant at the time (Table 1). The VE against severe disease of the mRNA-1273.815 vaccine was assumed to be the same as the booster VE against severe disease for the Moderna bivalent Autumn 2022 vaccine as estimated in the Kaiser Permanente prospective cohort study by Tseng et al. [36]. This study estimated the effectiveness of the Moderna bivalent booster for severe disease only; therefore, the VE for infection was assumed to be the same as for the Moderna monovalent booster against BA.1/2 [37]. The relative vaccine effectiveness (rVE) rates estimated by Kopel et al. [18] between the Moderna and Pfizer-BioNTech bivalent vaccines were used to estimate the VEs against severe disease and infection for the XBB.1.5 BNT162b2 vaccine. The rVE against hospitalisation was used as a proxy for severe disease. Because there were no data on infections, the rVE against outpatient visit was used as a proxy for infection. Waning was assumed to be equivalent to that of the monovalent vaccines against BA.1/BA.2 [38].

**Table 1.** Comparative vaccine effectiveness of the updated mRNA vaccines.

|  | Infection (%) |  | Severe Disease (%) |  |
| --- | --- | --- | --- | --- |
|  | VE | Waning | VE | Waning |
| mRNA-1273.815 vaccine | 57.1 | 4.8 | 84.3 | 1.4 |
| XBB.1.5 BNT162b2 vaccine | 54.8 | 4.8 | 82.6 | 1.4 |

### Model calibration and projection

We performed a manual model calibration to estimate the transmissibility parameter to reflect the pandemic history experienced in Germany from February 2020 through August 2023. To reflect the changing nature of the reporting on infections over time, we chose two different calibration targets for the daily number of SARS-CoV-2 infections. Between 4 February 2020 and 15 April 2022, the calibration target was the daily incidence of reported COVID-19 cases, representing both symptomatic and asymptomatic infections, as reported by the Institute for Health Metrics & Evaluation (IHME) [39]. Estimates from the IHME were chosen because they account for common biases that can result in underreporting of infections as described previously.

Although the IHME continued to report estimates until November 2022, estimates of infections began to diverge from reported estimates from RKI. Between 15 May 2022 and 15 April 2023, the calibration target was the daily number of reported cases from RKI. Because the SEIR model estimates the total number of infections, including asymptomatic infections, the number of reported cases was adjusted, accounting for the proportion of SARS-CoV-2 infections that are asymptomatic using a meta-analysis by Shang et al. [40]. For the period between 15 April 2022 and 15 May 2022, a linear interpolation between the two calibration targets was assumed to avoid sudden decreases in the number of infections that could cause instability during the calibration processes.

The fit of the model calibration to the target data was assessed in a qualitative way by visually comparing the daily case incidence predicted by the model to that for IHME during the period between 4 February 2020 and 15 April 2022, and by comparing to estimates by RKI between the period between 15 May 2022 and 15 April 2023 (Figure 1). Additional details about model calibration are described in the Technical Appendix.

**Figure 1.**
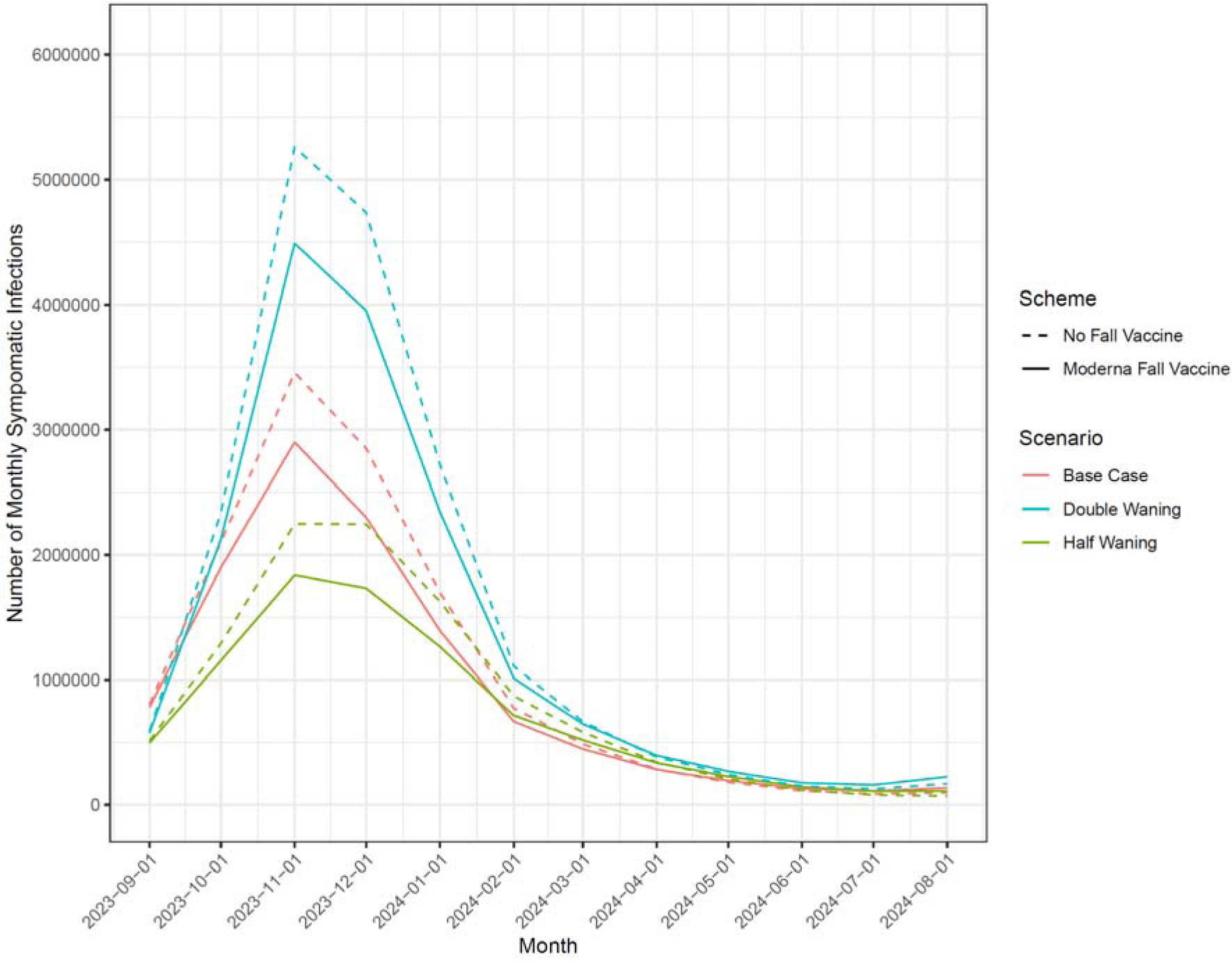
Incidence of symptomatic SARS-CoV-2 infection with and without mRNA-1273.815 Vaccine, by alternative assumptions of vaccine effectiveness waning and month.

For the time period of the actual analysis (projection) over Autumn 2023, the transmissibility parameters were chosen (a) to match the number of hospitalisations in the prior year (September 2022--August 2023) and (b) to reflect the typical seasonality pattern of acute respiratory infections with an increase of cases over the autumn, a peak in December, and a decrease in cases around spring. The actual values of the transmissibility parameter can be found in the Technical Appendix. All SEIR model analyses were conducted in R version 4.2.2 [41].

### Alternative infection scenarios

In addition to the base case scenario described above, additional calibrations were performed to allow the proportion of individuals with natural immunity and relative amount of residual VE against infection and hospitalisation to vary prior to the start of the analytic time period. Similar to that described in Kohli et al. [29], the waning rate of natural protection from SARS-CoV-2 infection was varied and the model was recalibrated to the same level as the base case.

### Cost-consequences model

The cost-consequences model consists of a decision tree using the number of infections from the dynamic model described above as input [29]. The number of asymptomatic (32.4%) and symptomatic (67.6%) cases [43], as well as the number of cases receiving outpatient care or hospitalised, is calculated from the overall number of infections. Hospitalized patients may receive regular hospital care, treatment in an intensive care unit, or ventilation, and hospitalisation rates are reduced depending on the vaccination status. Only in-hospital mortality is included in the model. Hospitalized patients are assumed to have received outpatient care before being hospitalised.

The treatment costs associated with the above disease states are reported in Table 2. Costs of each vaccination campaign were estimated to be €119/dose, including an administration fee for both mRNA vaccines (see Table 2). Indirect costs of vaccination have been included as 0.18 workdays of lost productivity. Indirect costs in the form of productivity and work losses were calculated using the duration of the different disease states and the average age-specific workforce participation and daily wage. A friction-cost approach was applied for COVID-19-related deaths using a friction period of 127 days [42]. QALYs have been used to capture the outcomes of the vaccination campaign. No disutility has been assumed for asymptomatic cases and no difference in disutilities was assumed between symptomatic cases with and without outpatient care. Detailed information on the values used can be found in Tables 3 and 4 and in the Technical Appendix.

**Table 2.** Resource utilization and cost parameters.

|  |  |  |  |  |  |
| --- | --- | --- | --- | --- | --- |
| Non-Hospitalised Care |  |  |  |  |  |
| Proportion seeking outpatient care<sup>a</sup> (%) |  |  | 15.8% |  |  |
| Hospitalisation Rates (%) |  |  |  |  |  |
| Age (years) | Unvaccinated<sup>b</sup> | No ICU or ventilation<sup>c</sup> | ICU only<sup>c</sup> | ICU with ventilation<sup>c</sup> | Mortality (overall)<sup>d</sup> |
| 0-4 | 1.3 | 92.9 | 0.0 | 7.1 | 0.0 |
| 5-17 | 0.5 | 77.2 | 21.2 | 1.6 | 0.1 |
| 18-29 | 0.7 | 84.7 | 8.1 | 7.2 | 0.1 |
| 30-39 | 1.2 | 77.2 | 9.6 | 13.1 | 0.2 |
| 40-49 | 1.6 | 69.8 | 11.2 | 19.1 | 0.5 |
| 50-59 | 1.7 | 69.8 | 11.2 | 19.1 | 2.4 |
| 60-69 | 8.4 | 59.2 | 12.6 | 28.3 | 1.9 |
| 70-79 | 11.1 | 59.2 | 12.6 | 28.3 | 6.6 |
| ≥80 | 16.9 | 73.2 | 13.5 | 13.3 | 7.0 |
| Endemic vaccination-related costs<sup>e</sup> |  |  | €119 |  |  |
| Cost of outpatient care<sup>f</sup> |  |  | €122.02 |  |  |
| Hospitalisation costs, by level of care [52] |  |  |  |  |  |
| No ICU or ventilation<sup>c</sup> |  |  | €6,900 |  |  |
| ICU only<sup>c</sup> |  |  | €27,981 |  |  |
| ICU with ventilation<sup>c</sup> |  |  | €38,500 |  |  |
| Hospitalisation recovery cost<sup>g</sup> |  |  | €15.25 |  |  |

<sup>a</sup>weighted average calculated from Robert Koch Institut [53]
<sup>b</sup>Data represent 4th quarter of 2020 (to capture unvaccinated); estimates adjusted for Omicron [51,53,54]
<sup>c</sup>Intensive care unit
<sup>d</sup>Data represent 1st Quarter of 2023 (reflects Omicron [2])
<sup>e</sup>Assumption (Vaccine €109 price to patient; Administration fee €10 [55]. As price for XBB.1.5 BNT162b2 was not available, same price as for mRNA-1273.815 was assumed)
<sup>f</sup>Assumes 1.6 outpatient visits at a cost of €76.26 per visit [56,57]
<sup>g</sup>Assumes 0.2 outpatient visits at a cost of €76.26 per visit [56,57]

**Table 3.** Utility values and quality-adjusted life years (QALY) decrement due to events in the consequences decision tree.

| Location of Care | Disutility | Duration | QALY Decrement | Source(s) |
| --- | --- | --- | --- | --- |
| Short-Term Infection Period |  |  |  |  |
| Not hospitalised | 0.59 | 4.29 days | 0.008 | [56,58,59] |
| Hospitalised, no ICU or Ventilator | 0.425 | 9 days | 0.010 | [56,59-61] |
| Hospitalised, ICU | 0.425 | 11.8 days | 0.014 |  |
| Hospitalised, ICU with ventilator | 0.425 | 25.2 days | 0.029 |  |
| Recovery From Hospitalisation During the Short-Term Infection Period |  |  |  |  |
| No ICU or ventilator | 0.0 | 0.0 | 0.00 | Assumption* |
| ICU | 0.0 | 0.0 | 0.00 |  |
| ICU with ventilator | 0.0 | 0.0 | 0.00 |  |
| Post-Infection |  |  |  |  |
| Not hospitalised |  |  | 0.028 | [62] |
| Hospitalised |  |  | 0.122 | [63] |
| Lifetime QALY loss due to mortality, discounted at 3% |  |  |  |  |
| 0-4 years |  |  | 29.46 |  |
| 5-17 years |  |  | 28.29 |  |
| 18-29 years |  |  | 26.10 |  |
| 30-39 years |  |  | 23.45 |  |
| 40-49 years |  |  | 20.37 |  |
| 50-59 years |  |  | 16.71 |  |
| 60-69 years |  |  | 12.65 |  |
| 70-79 years |  |  | 8.40 |  |
| ≥80 years |  |  | 3.14 |  |

ICU: intensive care unit
\*All short-term quality-of-life impacts are assumed to be captured by the short-term infection period QALY decrements

**Table 4.** Productivity and work loss parameters used in the model.

| Model Parameter | Value | Source |
| --- | --- | --- |
| Labor Force Participation Rates |  |  |
| 0-4 years | 0.0% | [64] |
| 5-17 years | 2.9% |  |
| 18-29 years | 69.4% |  |
| 30-39 years | 83.3% |  |
| 40-49 years | 86.4% |  |
| 50-59 years | 76.2% |  |
| 60-69 years | 47.7% |  |
| 70-79 years | 7.34% |  |
| ≥80 years | 0.0% |  |
| Average Daily Wage (€) |  |  |
| 0-4 years | 0.00 | [65] |
| 5-17 years | 17.30 |  |
| 18-29 years | 101.50 |  |
| 30-39 years | 163.20 |  |
| 40-49 years | 175.10 |  |
| 50-59 years | 174.20 |  |
| 60-69 years | 60.30 |  |
| 70-79 years | 59.10 |  |
| ≥80 years | 56.50 |  |
| Workdays Lost for: |  |  |
| Vaccine Administration* | 0.18 | [66] |
| Not Hospitalised | 4.29 | [56]<br>6 days with adjustment for weekend (multiplied by 5/7) |
| Hospitalised |  |  |
| No ICU or ventilator | 9 | [61] |
| ICU only | 11.8 | [61] |
| Ventilator | 25.2 | [60] |
| Hospitalisation recovery | 0 | Assumption (no data available) |

ICU, intensive care unit; IQWiG, Institute for Quality and Efficiency in Health Care; US, United States
\*Based on US data. Assumed that 30% of patients would receive the vaccine in the pharmacy, corresponding to 0.2 hours of time lost, and the remaining 70% of patients would receive the vaccine in the physician's office, corresponding to 2 hours of time lost [66]. Assuming an 8-hour workday, 0.18 total workdays would be lost for vaccine administration.
Discount rate of 3% (Version 7 of the IQWiG general guidelines [45])
Life tables from Federal Statistical Office

The results of the cost-consequences model are reported as ICER and as ROI and BCR for cost-saving scenarios. ROI has been calculated by dividing the health insurance cost-savings in COVID-19-related costs by the costs of the vaccination campaign and the social ROI included indirect costs, dividing the societal cost-savings by the vaccination costs [43]. For the BCR, the QALY gains have been transformed into monetary value using a WTP threshold of €50,000 per QALY gained and added to the COVID-19-related costs before dividing by the costs of the vaccination campaign.

The base-case economic analyses were conducted using a healthcare payer perspective. A scenario analysis was conducted using the societal perspective, which also included lost productivity costs [44]. All costs were adjusted to reflect 2022 Euros. Because the time horizon for the analytical period is only 1 year, none of the costs was discounted and QALY losses due to COVID-19-related mortality were discounted at 3% [45].

### Sensitivity Analysis

Deterministic sensitivity analyses (DSAs) were performed on epidemiological and economic parameters. For the DSAs, the percentage with symptoms, hospitalisation rates in the unvaccinated, hospitalisation level of care, and in-hospital mortality rates were varied according to their 95% confidence intervals (CIs). All other inputs were varied by +/-20% of the base-case value.

## RESULTS

### mRNA-1273.815 campaign vs no vaccination

In base-case simulations covering the timeframe of 1 September 2023 to 31 August 2024, the mRNA-1273.815 campaign was found to be associated with lower costs and better clinical outcomes (as measured by infection incidence, hospitalisations, and deaths) compared with no vaccination.

In the base case, the model predicted there will be 12,930,549 symptomatic infections, 305,711 hospitalisations, and 11,125 deaths between 1 September 2023 and 31 August 2024 if no COVID-19 vaccine is introduced. If the mRNA-1273.815 campaign is given to adults aged 60 years and older, in addition to at-risk adults aged 30-59 years, 1,697,851 symptomatic infections, 85,439 hospitalizations, and 4,057 deaths will be prevented corresponding to a 13.1%, 27.9%, and 36.8% reduction respectively. Similar magnitudes of reduction were exhibited when the assumed waning of VE was varied by 50% and 200% of the base case (Figure 1).

The cost-effectiveness analysis showed that the mRNA-1273.815 campaign was dominant in the societal and healthcare payer base cases (Table 5). In the DSAs, the mRNA-1273.815 campaign remained dominant in the societal perspective except in the case where the VE against infection was at the lower bound of its 95% CI (ICER €3,357/QALY gained). From the healthcare payer perspective, the mRNA-1273.815 campaign was dominant in most cases and was highly cost-effective (ICERs ranging from €12 to €5,375 per QALY gained) when assuming lower initial VE, or faster VE waning, against hospitalisation or infection (see Table 5, Technical Appendix). In the probabilistic sensitivity analysis, the mRNA-1273.815 campaign also was found to be dominant in 46% of all simulations and cost-effective with an ICER of less than €1,500 per QALY gained in 100% of the simulations.

**Table 5.** Base-case cost-effectiveness of mRNA-1273.815 campaign vs no vaccination (healthcare payer perspective)

| Vaccination Strategy | Costs (€) | QALYs Lost | Δ Costs (€) | Δ QALYs Gained | ICER (Δ Cost/QALY Gained) |
| --- | --- | --- | --- | --- | --- |
| mRNA-1273.815 campaign | 5,079,554,822 | 484,679 | -- | -- |  |
| No vaccination | 5,126,177,326 | 581,617 | 46,622,504 | -96,938 | mRNA-1273.815 Dominant |

ICER, incremental cost-effectiveness ratio; QALY, quality-adjusted life-year

### Return on investment analysis

As vaccination compared with no vaccination was dominant in most analyses, separate sensitivity analyses were undertaken to assess how ROI, from both healthcare payer and societal perspectives, for the mRNA-1273.815 campaign would vary with respect to variations in key model parameters compared to no vaccination. societal ROI was approximately 1.39 whereas, from the healthcare payer perspective, the base-case ROI was 1.04. From a societal perspective this finding was moderately to highly insensitive to variation in model parameters, indicating cost savings with the exception of VE against infection where the range crossed 1.0 (Figure 2). ROI from the payer perspective exhibited less overall sensitivity to variation in model parameters, but ranges crossed 1.0 for some key parameters including VE against infection and hospitalisation, Moderna vaccine price, and cost/rate of hospitalisation (Figure 9, Technical Appendix). The corresponding BCRs are 4.51 and 4.73 from a payer and societal perspective, respectively, and assuming a WTP threshold of €50,000 per QALY gained (Figure 9, Technical Appendix).

**Figure 2.**
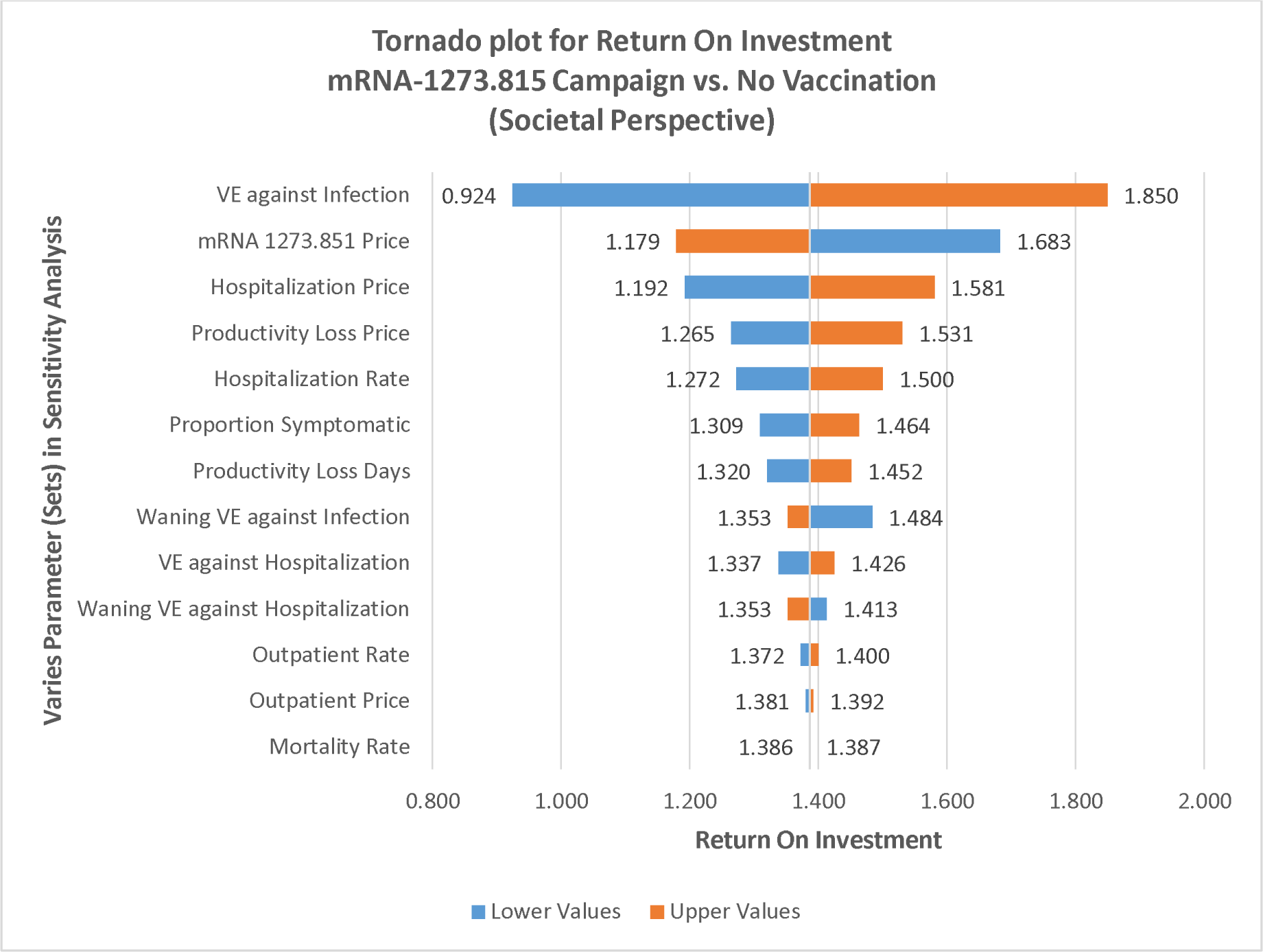
Deterministic sensitivity analyses of societal return on investment.

### Comparative analysis of mRNA-1273.815 vs XBB.1.5 BNT162b2 campaigns

In base-case simulations covering the time frame of 1 September 2023--31 August 2024, the mRNA-1273.815 campaign was found to be associated with lower costs and better clinical outcomes (as measured by infection incidence, hospitalisations, and deaths), compared with the XBB.1.5 BNT162b2 campaign. In the base case, the model predicted that vaccination with the mRNA-1273.815 campaign would result in 90,123 fewer symptomatic infections, 3,471 hospitalisations, and 157 deaths compared to vaccination with the XBB.1.5 BNT162b2 campaign (Table 6).

**Table 6.** Base-case cost-effectiveness of mRNA-1273.815 campaign vs XBB.1.5 BNT1262b2 campaign (healthcare payer perspective)

| Vaccination Strategy | Costs (€) | QALYs Lost | Δ Costs (€) | Δ QALYs Gained | ICER (Δ Cost/QALY Gained) |
| --- | --- | --- | --- | --- | --- |
| mRNA-1273.815 campaign | 5,079,554,822 | 484,679 | -- | -- |  |
| XBB.1.5 BNT1262b2 campaign | 5,138,327,691 | 489,356 | 58,772,869 | -4,677 | mRNA-1273.815 Dominant |

ICER, incremental cost-effectiveness ratio; QALY, quality-adjusted life-year

From a cost-effectiveness perspective, the mRNA-1273.815 campaign was dominant in the societal and healthcare payer base cases. In the base-case analysis, reflecting the healthcare payer perspective, the mRNA-1273.815 vaccine had greater initial VE than the XBB.1.5 BNT162b2 vaccine. Consequently, considering also the assumption of equivalent endemic vaccination costs, the mRNA-1273.815 campaign was estimated to yield lower total direct medical and indirect costs, as well as fewer QALYs lost due to symptomatic infection, than the XBB.1.5 BNT162b2 campaign. The robustness of these findings with respect to variations in a wide variety of model parameters was demonstrated in sensitivity analyses (see Technical Appendix).

### Value-based price premium of mRNA-1273.815 vs XBB.1.5 BNT162b2 vaccine

As noted previously, the base case assumes equivalent endemic vaccination costs between the Autumn 2023 mRNA-1273.815 and XBB.1.5 BNT162b2 vaccines. However, to the extent that the higher initial VE of mRNA-1273.815 translates into a relative reduction in infection incidence – which then is associated with lower direct medical and indirect costs as well as fewer QALYs lost due to infection – a higher price for mRNA-1273.815 would be economically justifiable. The analysis to determine potential value-based price premiums for mRNA-1273.815 vs XBB.1.5 BNT162b2 at various WTP thresholds and alternative rVE assumptions is shown in Figure 3.

**Figure 3.**
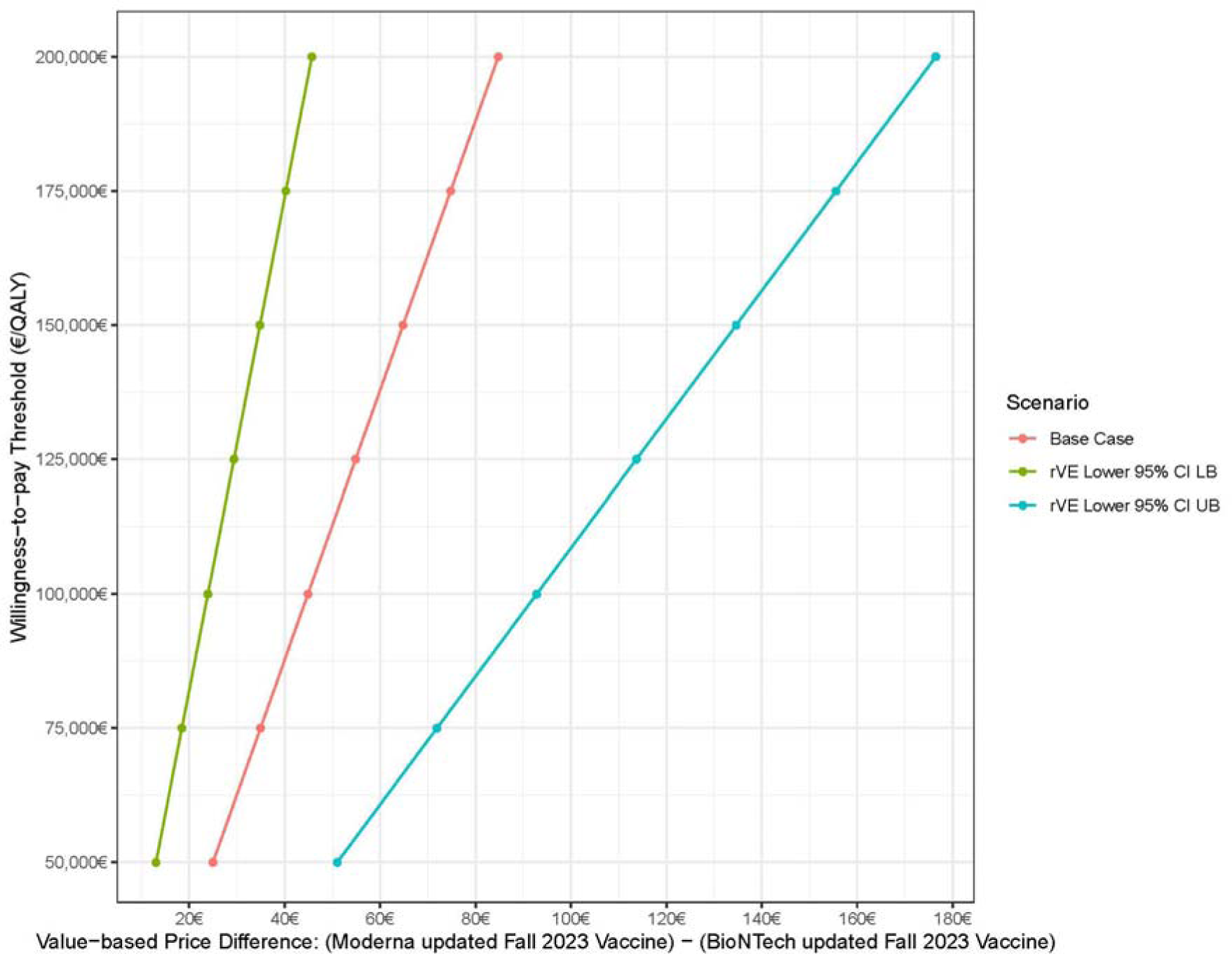
Value-based price premium (mRNA-1273-815 vs XBB.1.5 BNT1262b2 vaccine) at various willingness-to-pay thresholds, by rVE scenario.

At the threshold of €50,000 per QALY gained, considered reasonable value for money in Germany, the value-based price premium was €24.95, a nearly 23% premium (range €13.00-51.00). As the WTP threshold increases, the value-based price favoring the mRNA-1273.815 vaccine also increases: at thresholds of €100,000 and €150,000 per QALY gained, respectively, the price premiums favoring the mRNA-1273.815 vaccine are estimated to be €44.88 (range €23.90--€92.82) and €64.82 (range €34.80--€134.64).

## DISCUSSION

In this analysis we quantified the clinical and economic impact of an Autumn mRNA-1273.815 campaign compared with no Autumn vaccination and to an Autumn XBB.1.5 BNT162b2 campaign. We found that, compared with no vaccination, the mRNA-1273.815 campaign is predicted to prevent 1,697,851 symptomatic infections, 85,439 hospitalisations, and 4,057 deaths. Compared with an XBB.1.5 BNT162b2 campaign, the mRNA-1273.815 campaign is predicted to prevent 90,123 symptomatic infections, 3,471 hospitalisations, and 157 deaths. When considering cost-effectiveness, in base-case analyses we found the mRNA-1273.815 campaign dominated both the no vaccination and the XBB.1.5 BNT162b2 campaign.

In a meta-analysis of studies of the monovalent [17,19-21] and bivalent versions [18] in the general and immunocompromised populations, the Moderna mRNA COVID-19 vaccine (mRNA-1273) was found to be more effective than the Pfizer-BioNTech mRNA COVID-19 vaccine (BNT162b2). We have shown that the mRNA COVID-19 vaccines also differ, not only in their characteristics and compositions, but in their health economic performance. Assuming that the VE for mRNA-1273.815 is greater than for XBB.1.5 BNT162b2, as observed with previous versions of the vaccines and variants, our cost-effectiveness analysis for the eligible population suggests that a price premium of at least €24.95 per dose could be economically justifiable assuming the base case rVE (mRNA-1273 vs BNT162b2: 5.1% infection; 9.8% hospitalisation) at a minimum WTP threshold of €50,000 per QALY gained.

As the COVID-19 pandemic begins to shift to the endemic phase, the future epidemiology of COVID-19 is highly uncertain. Understanding time trends in COVID-19 disease is additionally complicated by temporal changes in reporting and across regions. To understand the impact of different incidence scenarios, we considered two additional incidence scenarios, varying the waning rate of natural immunity after infection during the B.A.4/5 period. Across all scenarios, the mRNA-1273.815 campaign is predicted either to be dominant or highly cost-effective (€904 per QALY gained).

In addition, understanding how variants will evolve, and the subsequent impact on immune-or vaccine-mediated protection, is challenging. For this analysis we assumed VE against hospitalization is well-matched to the circulating variant using data from the bivalent vaccine, but the true VE against infection and hospitalization, and the associated waning of protection, are unknown. Across a range of VE and waning scenarios we found the mRNA-1273.815 campaign to be dominant, or highly cost effective (less than €5,500 per QALY gained), relative to no vaccination or to the XBB.1.5 BNT162b2 campaign.

The modelling analyses are subject to simplifying assumptions on population dynamics and pathogen characteristics that could impact the generalizability of our results. First, due to the complexity of COVID-19 disease over the course of the pandemic, we are unable to capture prior infection history in the model which may result in an underestimation of the level of residual protection at the start of the analyses. Next, we use a contact matrix developed prior to COVID-19 and adjust this base matrix using information on masking and social distancing practices. These two metrics might not capture the complexity of human mobility and behavior observed during the pandemic which could lead to incorrect mixing patterns and thus impact the effect of vaccination. Further, our calibration targets are subject to many errors due to changes in testing and reporting practices, and disease severity over time. A secondary calibration target of COVID-19 hospitalisations in the prior year (September 2022--August 2023), a more stable metric that is less prone to reporting biases, was used to minimize the impact of these biases on model projections.

In addition, other data that may increase the value of vaccination also were not included in the analysis. For example, post-infection costs and QALY decrements were only applied for a limited duration. Additionally, although analyses from the societal perspective were included, the emphasis was on short-term lost productivity for the COVID-19 patient, whereas the broader aspects of lost productivity, such as caregiver time and the impact of post- and/or long-COVID, are not included, nor are the consequences of weighing utility losses associated with severe disease higher than non-severe disease and the associated utility losses for families [46,47].

To the best of our knowledge, ours is the first study evaluating a COVID-19 vaccination program from a health economic perspective in the German setting. Quantifying the economic burden of COVID-19 is an essential consideration for evaluating the value of therapeutic and preventive interventions against COVID-19 disease [26]. STIKO also uses mathematical modelling as well as health economics in their Standard Operating Procedures for vaccination recommendation decision making [48].

Overall, we have demonstrated that vaccination with the Moderna updated COVID-19 vaccine represents a cost-saving measure from both a healthcare payer and a societal perspective compared with no vaccination. This finding is consistent with systematic literature reviews of economic evaluations of COVID-19 vaccination strategies, which provide evidence that COVID-19 vaccination strategies are economically favourable [49-51].

## CONCLUSION

We have shown that COVID-19 vaccination results in lower healthcare costs and, assuming that the mRNA-1273.815 vaccine is more effective than XBB.1.5 BNT162b2, cost savings. Further, from a healthcare payer perspective, mRNA-1273.815 also represents good value for money across a broad range of scenarios considering a widely accepted cost per QALY threshold compared with both no vaccination and to XBB.1.5 BNT162b2. For these reasons a portfolio of vaccines is essential in addition to reasons such as maintaining supply, patient-individual care, a doctor’s freedom of therapy, and competition as seen in the COVID-19 vaccine market, and thus a portfolio of vaccines should also be available in the evolving endemic.

## Supporting information

Technical Appendix

## Data Availability

All data produced in the present study are available upon reasonable request to the authors.

## ACKNOWLEDGEMENTS AND DISCLOSURES

Medical writing and editorial assistance were provided by K Ian Johnson BSc, MBPS, SRPharmS of MEDiSTRAVA, in accordance with Good Publication Practice (GPP) guidelines, funded by Moderna, Inc., and under the direction of the authors.

## CONFLICT OF INTEREST

EB, KJ, SS, BU, and NVV are employees of Moderna and may hold stock/stock options in the company. MK is a shareholder in Quadrant Health Economics Inc, which was contracted by Moderna, Inc., to conduct this study. KF, AL, and MM are consultants at Quadrant Health Economics Inc.

## AUTHOR CONTRIBUTIONS

AL, EB, KF, KJ, MK, MM, SS, BU, and NVV were involved in study design and interpretation of the analysis. KJ and SS programmed the model and conducted the analysis. EB, KJ, SS, and BU contributed to development of the initial draft of the manuscript, and all remaining co-authors critically revised the manuscript and approved the final version.

## FUNDING

This study was funded by Moderna, Inc. Employees of Moderna participated in the design and conduct of the study; collection, management, analysis, and interpretation of the data; preparation, review, or approval of the manuscript; or the decision to submit the manuscript for publication.

## Notes

### Author Declarations

Source data are fully detailed in the references in the manuscript and supplementary files. All have been obtained from published sources.

## REFERENCE LIST

[1] UN. WHO chief declares end to COVID-19 as a global health emergency United Nations2023 [13th September 2023]. Available from: https://news.un.org/en/story/2023/05/1136367

[2] RKI. COVID-19-Todesfälle in Deutschland. Robert Koch-Institut. 2023.

[3] RKI. Decision on the implementation of the COVID-19 vaccination into the general recommendations of the STIKO 2023. Robert Koch Institute: Standing Committee on Vaccination (Ständigen Impfkommission, STIKO)/. 2023.

[4] Vygen-Bonnet S, Koch J, Bogdan C, et al.: Beschluss und Wissenschaftliche Begründung der Ständigen Impfkommission (STIKO) für die COVID-19-Impfempfehlung. Epidemiological Bulletin. 2021;2:3–63.

[5] Koch J, Piechotta V, Berner R, et al. Empfehlung der STIKO zur Implementierung der COVID-19-Impfung in die Empfehlungen der STIKO 2023 und die dazugehörige wissenschaftliche Begründung. Epidemiological Bulletin. 2023;21:7–48.

[6] Link-Gelles R, Ciesla AA, Roper LE, et al. Early estimates of bivalent mRNA booster dose vaccine effectiveness in preventing symptomatic SARS-CoV-2 infection attributable to Omicron BA.5- and XBB/XBB.1.5-related sublineages among immunocompetent adults - increasing community access to testing program, United States, December 2022-January 2023. MMWR Morb Mortal Wkly Rep. 2023 Feb 3;72(5):119-124.

[7] Link-Gelles R, Weber ZA, Reese SE, et al. Estimates of bivalent mRNA vaccine durability in preventing COVID-19-associated hospitalization and critical illness among adults with and without immunocompromising conditions - VISION Network, September 2022-April 2023. Am J Transplant. 2023 Jul;23(7):1062-1076.

[8] WHO. Statement on the antigen composition of COVID-19 vaccines. World Health Organization. 2023.

[9] EC. Union Register of medicinal products for human use. European Commission. 2023. Accessed at https://ec.europa.eu/health/documents/community-register/html/h1507.htm

[10] Chalkias S, McGhee N, Whatley JL, et al. Safety and Immunogenicity of XBB.1.5-Containing mRNA Vaccines. medRxiv. 2023. DOI 10.1101/2023.08.22.23293434.

[11] PEI. CoV-2 Development and Authorisation. In: Paul-Ehrlich Institut, Federal Institute for Vaccines and Biomedicines. 2023.

[12] EMA. Spikevax (previously COVID-19 Vaccine Moderna) for active immunisation to prevent COVID 19 caused by SARS-CoV-2 in individuals 6 months of age and older. European Medicines Agency. 2023.

[13] EMA. Comirnaty: EMA approval of adapted COVID-19 vaccine targeting Omicron XBB.1.5. European Medicines Agency. 2023.

[14] Ali K, Berman G, Zhou H, et al. Evaluation of mRNA-1273 SARS-CoV-2 vaccine in adolescents. N Engl J Med. 2021 Dec 9;385(24):2241-2251.

[15] Baden LR, El Sahly HM, Essink B, et al. Efficacy and safety of the mRNA-1273 SARS-CoV-2 vaccine. N Engl J Med. 2021 Feb 4;384(5):403–416.

[16] Polack FP, Thomas SJ, Kitchin N, et al. Safety and efficacy of the BNT162b2 mRNA Covid-19 vaccine. N Engl J Med. 2020 Dec 31;383(27):2603–2615.

[17] Hulme WJ, Horne EMF, Parker EPK, et al. Comparative effectiveness of BNT162b2 versus mRNA-1273 covid-19 vaccine boosting in England: matched cohort study in OpenSAFELY-TPP. BMJ. 2023 Mar 15;380:e072808.

[18] Kopel H, Nguyen VH, Boileau C. Comparative effectiveness of the bivalent (Original/Omicron BA.4/BA.5) mRNA COVID-19 vaccines mRNA-1273.222 and BNT162b2 bivalent in adults in the United States. medRxiv. 2023. DOI 10.1101/2023.07.12.23292576.

[19] Mayr FB, Talisa VB, Shaikh OS, et al. Comparative COVID-19 Vaccine effectiveness over time in veterans. Open Forum Infect Dis. 2022 Jul;9(7):ofac311.

[20] Nguyen VHBC, Bogdanov A. Relative effectiveness of BNT162b2, mRNA-1273, and Ad26.COV2.S vaccines and homologous boosting in preventing COVID-19 in adults in the US. medRxiv. 2023. DOI 10.1101/2023.02.10.23285603.

[21] Ono S, Michihata N, Yamana H, et al. Comparative effectiveness of BNT162b2 and mRNA-1273 booster dose after BNT162b2 primary vaccination against the Omicron variants: a retrospective cohort study using large-scale population-based registries in Japan. Clin Infect Dis. 2023 Jan 6;76(1):18–24.

[22] Wang X, Haeussler K, Spellman A, et al. Comparative effectiveness of mRNA-1273 and BNT162b2 COVID-19 vaccines in immunocompromised individuals: a systematic review and meta-analysis using the GRADE framework. Front Immunol. 2023;14:1204831.

[23] Bruxvoort KJ, Sy LS, Qian L, et al. Real-world effectiveness of the mRNA-1273 vaccine against COVID-19: Interim results from a prospective observational cohort study. Lancet Reg Health Am. 2022 Feb;6:100134.

[24] Florea A, Sy LS, Luo Y, et al. Durability of mRNA-1273 against COVID-19 in the time of Delta: Interim results from an observational cohort study. PLoS One. 2022;17(4):e0267824.

[25] Tenforde MW, Patel MM, Gaglani M, et al. Effectiveness of a third dose of Pfizer-BioNTech and Moderna vaccines in preventing COVID-19 hospitalization among immunocompetent and immunocompromised adults - United States, August-December 2021. MMWR Morb Mortal Wkly Rep. 2022 Jan 28;71(4):118-124.

[26] Richards F, Kodjamanova P, Chen X, et al. Economic burden of COVID-19: a systematic review. Clinicoecon Outcomes Res. 2022;14:293–307.

[27] Padula WV, Malaviya S, Reid NM, et al. Economic value of vaccines to address the COVID-19 pandemic: a U.S. cost-effectiveness and budget impact analysis. J Med Econ. 2021 Jan-Dec;24(1):1060-1069.

[28] Utami AM, Rendrayani F, Khoiry QA, et al. Economic evaluation of COVID-19 vaccination: a systematic review. J Glob Health. 2023 Jan 14;13:06001.

[29] Kohli M, Maschio M, Joshi K, et al. The potential clinical impact and cost-effectiveness of the updated COVID-19 mRNA Fall 2023 vaccines in the United States. medRxiv. 2023. DOI 10.1101/2023.09.05.23295085.

[30] Shiri T, Evans M, Talarico CA, et al. Vaccinating adolescents and children significantly reduces COVID-19 morbidity and mortality across all ages: a population-based modeling study using the UK as an example. Vaccines (Basel). 2021 Oct 15;9(10).

[31] Mossong J, Hens N, Jit M, et al. Social contacts and mixing patterns relevant to the spread of infectious diseases. PLoS Med. 2008 Mar 25;5(3):e74.

[32] Holstiege J, Akmatov MK, Kohring C, et al. Patients at high risk for a severe clinical course of COVID-19 - small-area data in support of vaccination and other population-based interventions in Germany. BMC public health. 2021 Sep 28;21(1):1769.

[33] Hodcroft E. CoVariants: SARS-CoV-2 Mutations and Variants of Interest. Institute of Social and Preventive Medicine, University of Bern, Switzerland and SIB Swiss Institute of Bioinformatics, Switzerland. 2021.

[34] RKI. Impfquoten bei Erwachsenen in Deutschland. Robert Koch-Institut Epidemiologisches Bulletin. 2022 Jan 8; 49.

[35] PEI. Seasonal Influenza 2021/2022. Paul-Ehrlich-Institut, Federal Institute for Vaccines and Biomedicines. 2022. Accessed at https://www.pei.de/EN/medicinal-products/vaccines-human/influenza-flu/preseasons/influenza-flu-season-2021-2022-content.html

[36] Tseng HF, Ackerson BK, Sy LS, et al. mRNA-1273 bivalent (original and Omicron) COVID- 19 vaccine effectiveness against COVID-19 outcomes in the United States. Nat Commun. 2023 Sep 20;14(1):5851.

[37] Pratama NR, Wafa IA, Budi DS, et al. Effectiveness of COVID-19 vaccines against SARS-CoV-2 Omicron Variant (B.1.1.529): a systematic review with meta-analysis and meta-regression. Vaccines (Basel). 2022 Dec 19;10(12).

[38] Higdon MM, Baidya A, Walter KK, et al. Duration of effectiveness of vaccination against COVID-19 caused by the omicron variant. Lancet Infect Dis. 2022 Aug;22(8):1114–1116.

[39] IHME. COVID-19 Results Hans Rosling Center for Population Health, University of Washington, Seattle, US: Institute for Health Metrics and Evaluation. 2021. Accessed at https://covid19.healthdata.org/germany?view=cumulative-deaths&tab=trend

[40] Shang W, Kang L, Cao G, et al. Percentage of asymptomatic infections among SARS-CoV-2 Omicron variant-positive individuals: a systematic review and meta-analysis. Vaccines (Basel). 2022 Jun 30;10(7).

[41] R Foundation. R: A language and environment for statistical computing. R foundation for Statistical Computing Vienna, Austria 2023 [30th September 2023]. Accessed at https://www.R-project.org/

[42] Klaus A. Statistik der Bundesagentur für Arbeit, Grundlagen: Engpassanalyse – Methodische Weiterentwicklung. Bundesagentur für Arbeit Statistik/Arbeitsmarktberichterstattung. 2020.

[43] Sim SY, Watts E, Constenla D, et al. Return on investment from immunization against 10 pathogens in 94 low- and middle-income countries, 2011-2030. Health Aff (Millwood). 2020 Aug;39(8):1343-1353.

[44] RKI. Standardvorgehensweise (SOP) der Ständigen Impfkommission (STIKO) für die systematische Entwicklung von Impfempfehlunge. Robert Koch-Institut. 2018.

[45] IQWiG. General methods for the assessment of the relation of benefits to costs (Version 6.1). Institut für Qualität und Wirtschaftlichkeit im Gesundheitswesen. 2022 24 Jan.

[46] Lakdawalla DN, Doshi JA, Garrison LP, Jr., et al. defining elements of value in health care-a health economics approach: an ISPOR Special Task Force report [3]. Value Health. 2018 Feb;21(2):131–139.

[47] Postma M, Biundo E, Chicoye A, et al. Capturing the value of vaccination within health technology assessment and health economics: Country analysis and priority value concepts. Vaccine. 2022 Jun 26;40(30):3999–4007.

[48] STIKO. Standard Operating Procedure of the German Standing Committee on Vaccinations (STIKO) for the systematic development of vaccination recommendations. Robert Koch-Institut. 2018. Accessed at https://www.rki.de/EN/Content/infections/Vaccination/methodology/SOP.pdf?__blob=publicationFile

[49] Aidalina M, Khalsom S. COVID-19 vaccination: a systematic review of vaccination strategies based on economic evaluation studies. Med J Malaysia. 2023 May;78(3):411–420.

[50] Fu Y, Zhao J, Han P, et al. Cost-effectiveness of COVID-19 vaccination: A systematic review. J Evid Based Med. 2023 Jun;16(2):152–165.

[51] Zhou L, Yan W, Li S, et al. Cost-effectiveness of interventions for the prevention and control of COVID-19: Systematic review of 85 modelling studies. J Glob Health;12. 2022 Jun 15.

[52] Kraft K. COVID-19 Behandlung: Kosten pro Patient im fünfstelligen Bereich. PraxisVita: Barmer Ersatzkasse. 2020. Accessed at https://www.praxisvita.de/covid-19-behandlung-kosten-pro-patient-krankenkassen-veroeffentlichen-zahlen-19253.html

[53] RKI. COVID-19-Hospitalisierungen in Deutschland, Berlin: Zenodo. Robert Koch-Institut. 2023. Accessed at https://github.com/robert-koch-institut/COVID-19-Hospitalisierungen_in_Deutschland#covid-19-hospitalisierungen-in-deutschland

[54] Wang L, Berger NA, Kaelber DC, et al. COVID infection rates, clinical outcomes, and racial/ethnic and gender disparities before and after Omicron emerged in the US. medRxiv. 2022 Feb 22. DOI 10.1101/2022.02.21.22271300

[55] CGM. Lauer-Taxe® Online 4.0. Spikevax XBB1.5 Injektionsdispersion Einsdos-Fla.2023 Accessed at https://portal.cgmlauer.cgm.com/LF/default.aspx?p=12000

[56] Bilcke J, Coenen S, Beutels P. Influenza-like-illness and clinically diagnosed flu: disease burden, costs and quality of life for patients seeking ambulatory care or no professional care at all. PLoS One. 2014;9(7):e102634.

[57] KBV. Honorarbericht Quartal 4 2020. Kassenarztliche Bundesvereinigung. 2020. Accessed at https://www.kbv.de/media/sp/Honorarbericht_Quartal_4_2020.pdf.

[58] CDC. Isolation and Precautions for People with COVID-19. Centers for Disease Control and Prevention. 2023. Accessed at https://www.cdc.gov/coronavirus/2019-ncov/your-health/isolation.html.

[59] Hollmann M, Garin O, Galante M, et al. Impact of influenza on health-related quality of life among confirmed (H1N1)2009 patients. PLoS One. 2013;8(3):e60477.

[60] Karagiannidis C, Mostert C, Hentschker C, et al. Case characteristics, resource use, and outcomes of 101021 patients with COVID-19 admitted to 920 German hospitals: an observational study. Lancet Respir Med. 2020 Sep;8(9):853–862.

[61] Schilling J, Tolksdorf K, Marquis A, et al. The different periods of COVID-19 in Germany: a descriptive analysis from January 2020 to February 2021. Bundesgesundheitsblatt Gesundheitsforschung Gesundheitsschutz. 2021 Sep;64(9):1093–1106.

[62] Sandmann FG, Tessier E, Lacy J, et al. Long-term health-related quality of life in non-hospitalized coronavirus disease 2019 (COVID-19) cases with confirmed Severe Acute Respiratory Syndrome Coronavirus 2 (SARS-CoV-2) infection in England: longitudinal analysis and cross-sectional comparison with controls. Clin Infect Dis. 2022 Aug 24;75(1):e962–e973.

[63] PHOSP-COVID. Clinical characteristics with inflammation profiling of long COVID and association with 1-year recovery following hospitalisation in the UK: a prospective observational study. Lancet Respir Med. 2022 Aug;10(8):761–775.

[64] Bund-Laender Demografie Portal. Erwerbstaetigenquote nach Alter und Geschlecht, 2000 und 2020. 2023. Accessed at https://www.demografie-portal.de/DE/Fakten/erwerbstaetigenquote-alter.html.

[65] Destatis. Verdienststrukturerhebung. Niveau, Verteilung und Zusammensetzung der Verdienste und der Arbeitszeiten abhangiger Beschaftigungsverhaltnisse. Statistiches Bundesamt. 2020. Accessed at www.destatis.de/.

[66] Prosser LA, Harpaz R, Rose AM, et al. A cost-effectiveness analysis of vaccination for prevention of herpes zoster and related complications: input for national recommendations. Ann Intern Med. 2019 Mar 19;170(6):380–388.

