## Supplementary material for "Clinical impact and cost-effectiveness of the updated COVID-19 mRNA Autumn 2023 vaccines in Germany": Technical Appendix

### 1. SEIR Model Inputs: Burn-in Period (January 2020 to August 2023)

The model structure and underlying assumptions of the Susceptible-Exposed-Infected-Recovered (SEIR) model have previously been described in a Technical Appendix for the US mode [1].

The rate at which individuals move from the susceptible to exposed state depends on the force of infection, which is the function of the number of susceptible individuals in the population as well as the effective contact rate between susceptible and infected individuals. The force of infection among individuals in the different vaccination strata is reduced proportional to the average vaccine effectiveness in the population.

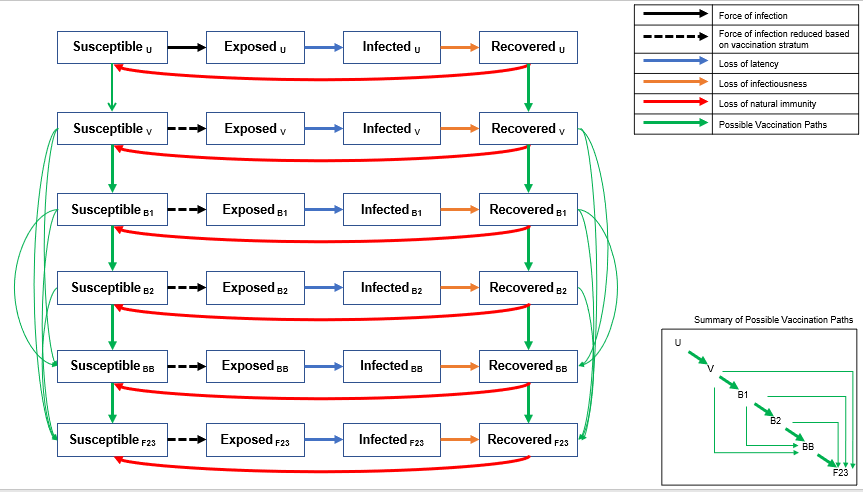

Source: Figure adapted from Kohli et al. 2023 [1]

For the burn-in period (January 2020 to August 2023), the variables that determine the rate of contact and the average vaccine effectiveness were estimated from data as described below. The estimation of other model transitions (movements into the Infected, Recovered, and Susceptible compartments) is described later in this section. The transmissibility of the virus changed over time as new variants emerged. The transmissibility was therefore estimated through a calibration process described below. Any inputs that change in the analytic period are described in section 2 below.

#### 1.1. Number of Susceptibles

All individuals in Germany were included in the model and were considered to be susceptible to COVID-19 infection at the start of the simulation (31 January 2020). The size of the German population in 2020 by age was obtained from the United Nations Department of Economic and Social Affairs [2].

Table 1. The model population size

| Age Group (Years) | Number |
| --- | --- |
| 0-9 | 7,749,369 |
| 10-19 | 7,625,944 |
| 20-29 | 9,593,301 |
| 30-39 | 10,821,918 |
| 40-49 | 10,108,496 |
| 50-59 | 13,389,921 |
| 60-69 | 10,619,715 |
| 70-79 | 7,500,977 |
| 80+ | 5,919,349 |
| **Total** | **83,328,988** |

#### 1.2. Number of Effective Contacts

The mixing patterns are based on contact matrices. As behaviours that impacted contact changed during the pandemic, the matrices are modified by a mobility index that accounts for social distancing and mask use.

##### 1.2.1. Mixing Patterns / Contact Matrices

Data on the age-specific mixing patterns in the general population for the Germany were obtained from Mossong et al. [3]. In the published contact matrices, the population was partitioned into 5-year age bands, and all individuals aged 75 years and older were grouped together. For this analysis, the age-specific mixing patterns were first converted into 10-year age bands.

We then assumed symmetry between the age groups (i.e., an effective contact between a person in age group *i* with a person from age group *j* is the same as an effective contact between a person in age group *j* with a person from age group *i),* weighted by the population estimates in age groups *i* and *j*.

Thus, $c_{ij}=\frac{1}{N_{j}}\times\frac{\left( c_{ij}^{*}\times N_{j} \right)+\left( c_{ji}^{*}\times N_{i} \right)}{2}$ ,

where:

$c_{ij}$ is the number of effective contacts between someone in age group *i* with someone from age group *j*

$c_{ij}^{*}$ is the number of effective contacts between someone in age group *i* with someone from age group *j* based on the original age-specific mixing patterns were first converted into 10-year age bands

$N_{i}$ is the population size in age group *i*.

The base contact matrix is presented in Table 2.

Table 2. Base contact matrix used in the model before applying scaling factors

| **Age Group of**  **Participant (Years)** | **Age Group of Contact (Years)** | | | | | | | | |
| --- | --- | --- | --- | --- | --- | --- | --- | --- | --- |
|  | **0-9** | **10-19** | **20-29** | **30-39** | **40-49** | **50-59** | **60-69** | **70-79** | **80+** |
| **0-9** | 2.5799 | 0.6484 | 0.6318 | 1.1516 | 0.5823 | 0.3800 | 0.2825 | 0.2870 | 0.3758 |
| **10-19** | 0.6381 | 4.7155 | 0.7986 | 0.6835 | 1.3539 | 0.3955 | 0.2499 | 0.1652 | 0.1632 |
| **20-29** | 0.7821 | 1.0046 | 3.3115 | 1.1405 | 1.1386 | 0.7986 | 0.4190 | 0.2976 | 0.2887 |
| **30-39** | 1.6081 | 0.9700 | 1.2866 | 2.3955 | 1.4628 | 0.8565 | 0.6917 | 0.6036 | 0.8591 |
| **40-49** | 0.7596 | 1.7946 | 1.1997 | 1.3663 | 2.2664 | 1.0404 | 0.7187 | 0.5289 | 0.6877 |
| **50-59** | 0.6565 | 0.6944 | 1.1146 | 1.0598 | 1.3781 | 1.9472 | 0.9706 | 0.7212 | 1.0842 |
| **60-69** | 0.3872 | 0.3480 | 0.4638 | 0.6788 | 0.7550 | 0.7698 | 1.5519 | 0.9362 | 0.5582 |
| **70-79** | 0.2778 | 0.1625 | 0.2327 | 0.4184 | 0.3925 | 0.4040 | 0.6612 | 1.1710 | 1.0763 |
| **80+** | 0.2871 | 0.1267 | 0.1781 | 0.4699 | 0.4027 | 0.4793 | 0.3112 | 0.8493 | 1.2977 |

Note: Since the population size of the age groups are not of equal size, the transformed contact matrix is not strictly symmetric.

During the pandemic, the rate of contact was reduced by behaviours such as social distancing and mask use. The magnitude of the impact is estimated as described in the next section.

##### 1.2.2. Social Distancing and Mask Use: Overall Scaling Factor

From 31 January 2020 to 24 June 2022, it was assumed that regular patterns of interaction were modified because of social distancing and use of masks to reduce effective contacts between individuals. Data on social distancing patterns and mask use were obtained from the Institute for Health Metrics and Evaluation (IHME) and used to adjust the base contact matrix based on these factors [4]. Daily estimates on social distancing patterns and mask use were obtained from the IHME for the time period 31 January 2020 through 24 June 2022, inclusive. For the time period after 31 July 2022, a single seasonality parameter was assumed to replace the social distancing patterns and mask use data. Between these two time points (24 June—31 July 2022), a linear interpolation between the scaling factors estimates was assumed to avoid an abrupt transition between these two methods.

For the base case, we assumed that mask use was 0% from 31 July 2022 onwards. We also assumed that social mobility would return to normal (baseline) after this date and stay at this level for the remaining time period. Mask use represents the percentage of the population who say they always wear a mask in public. Daily estimates of mask use were obtained from IHME [4]. It was assumed that 100% mask usage is associated with a 30% reduction in transmission and that a reduction in mask usage impacts the reduction in transmission proportionately [4]. Therefore, the following scaling factor (due to mask use) was applied daily to the base contact matrix:

$${Scaling Factor}_{Mask use}=1-\left( Mask use\left( \% \right)\times0.30 \right)$$

Daily estimates of the change in mobility were obtained from IHME [4] and was applied to reduce the number of contacts per person. Since age-specific data on changes in mobility were not available, the impact was applied equally to all age groups. Therefore, the following scaling factor (due to change in mobility) was applied daily to the base contact matrix:

$${Scaling Factor}_{Mobility}=1+Change in mobility (\%)$$

After July 31, 2022, a standard sinusoidal function was applied to vary the rate of contact by season [5]. The assumed peak was 15 February to account for greater time spent indoors in the winter. The assumed trough was 15 August to account for greater time spent outdoors in summer. The trough and peak were 95% and 105% of normal contact respectively [5].

We defined the seasonality function over time applied to the transmissibility parameter that the transition rate is a function of time $\beta\left( t \right)$ as follows:

$$Seasonality=\left( 1+\frac{\phi}{2}\sin\left( 2\pi t+\omega\right) \right)$$

where $\phi$ denotes the amplitude of seasonality (0.1 in the base case)

and $\omega$ is the phase shift of the sine function such that the peak was 15 February and the trough was 15 August.

The overall scaling factor was thus calculated as:

| Overall Scaling Factor | |  | | $\left[ {Scaling Factor}_{Mask use} \right] \times\left[ {Scaling Factor}_{Mobility} \right]$ | | from 30 January 2020 to 24 June 2022 |
| --- | --- | --- | --- | --- | --- | --- |
|  |  |  |  | Seasonality estimate | | after 31 July 2022 |
|  |  |  |  | Linear interpolation between above | | Between 24 June—31 July 2022 |
| Overall Scaling Factor |  | | $\left[ 1-\left( Mask use\left( \% \right)\times0.30 \right) \right] \times\left[ 1+Change in mobility (\%) \right]$ | | from 30 January 2020 to 24 June 2022 | |
|  |  |  | $\left( 1+\frac{\phi}{2}\sin\left( 2\pi t+\omega\right) \right)$ | | after 31 July 2022 | |
|  |  |  | Linear interpolation between above | | Between 24 June—31 July 2022 | |

The final monthly pattern for the scaling factor for both the burn-in period (January 2020 to July 2023) and the analysis period (September 2023 to August 2024) is shown in Figure 1.

Figure 1. Mobility scaling factor over time (burn-in and analysis period)

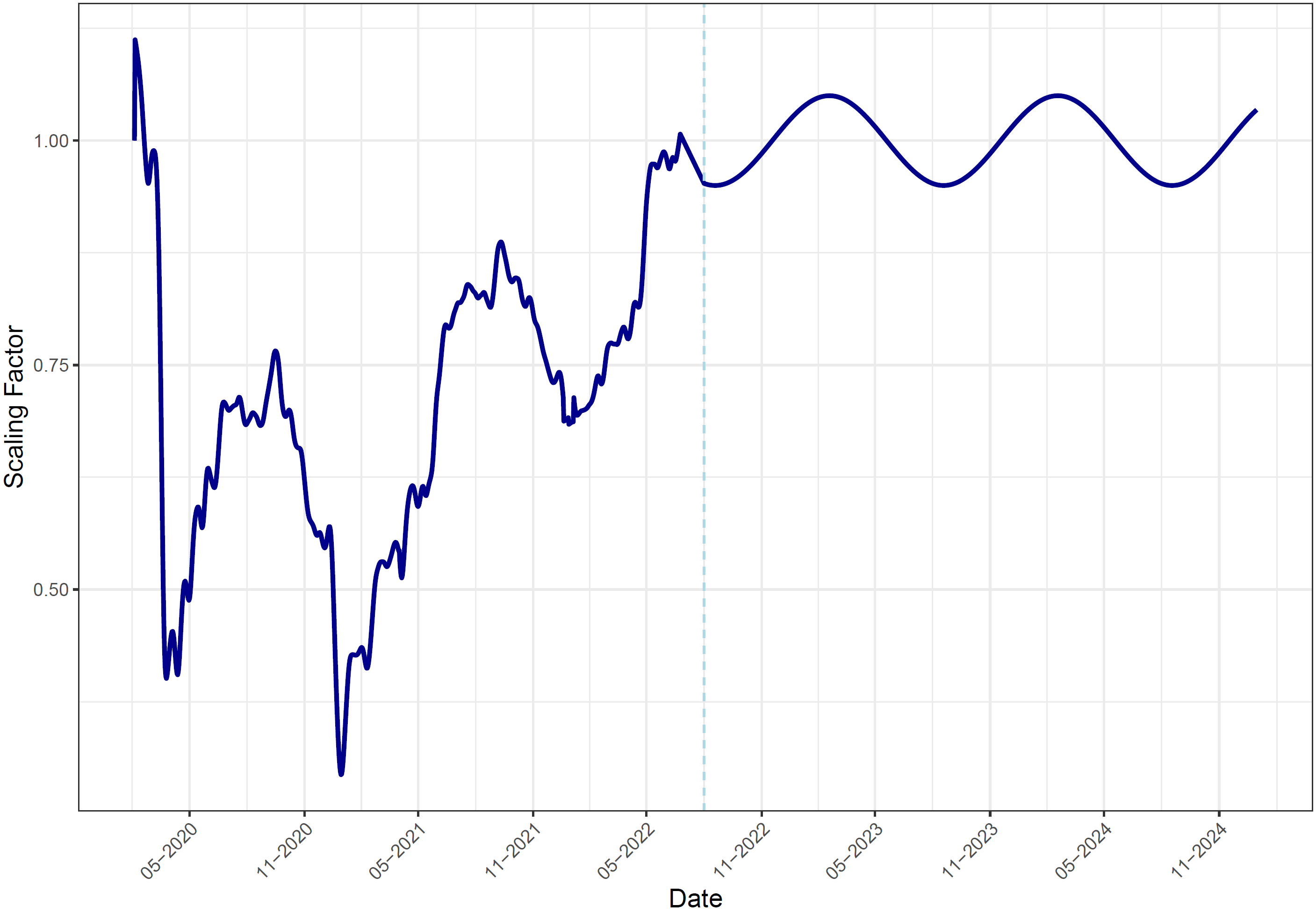

#### 1.3. Reduction in Effective Contacts Due to Vaccination

##### 1.3.1. Vaccine Coverage

A proportion of the population moves into the primary series and booster model strata according to vaccine uptake data from the Robert Koch Institute, as illustrated in Figure 2 to Figure 5.

Figure 2. Percentage of the population who have completed their primary series, by age group

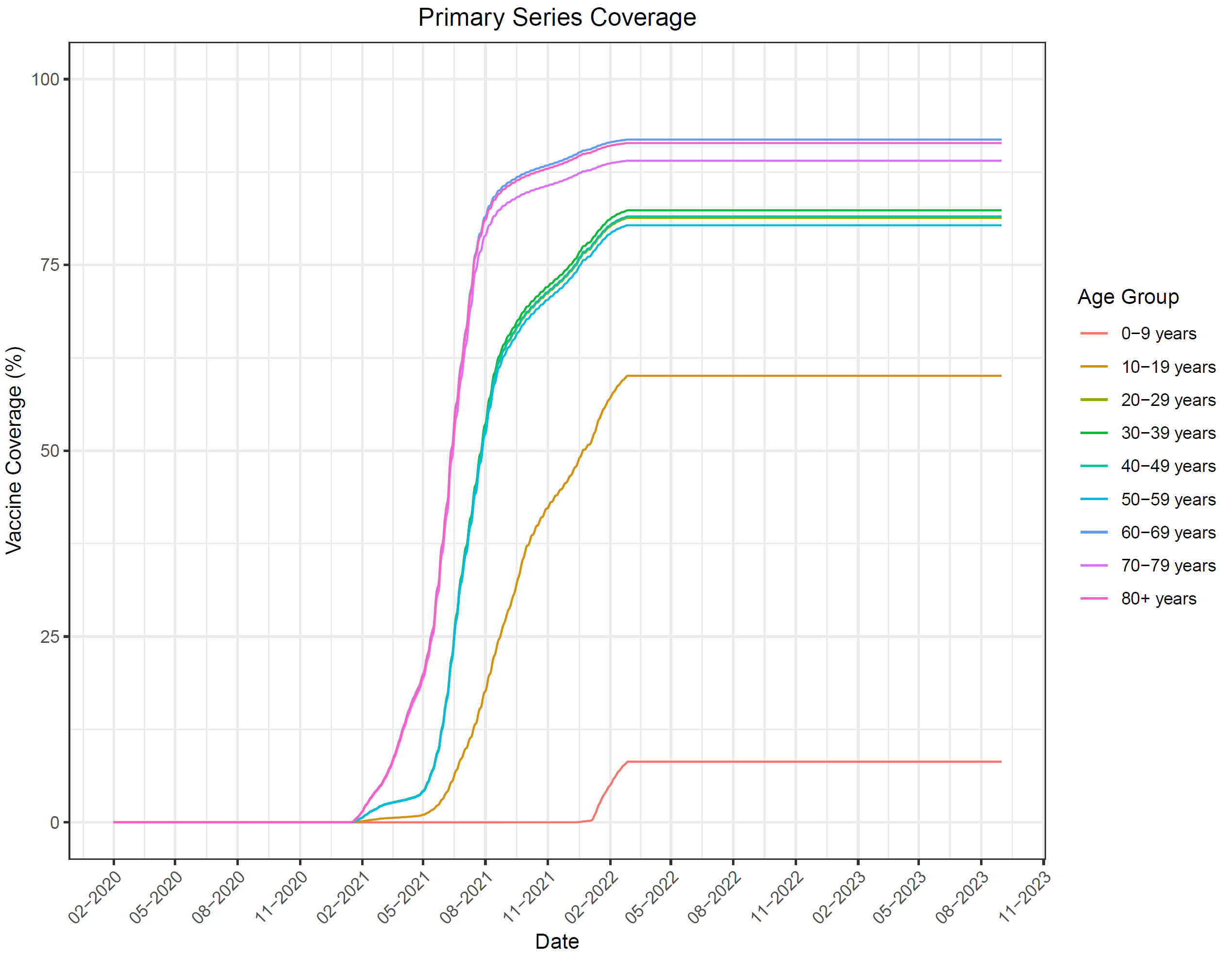

**Figure 3. Percentage of the population who have received a booster, by age group**

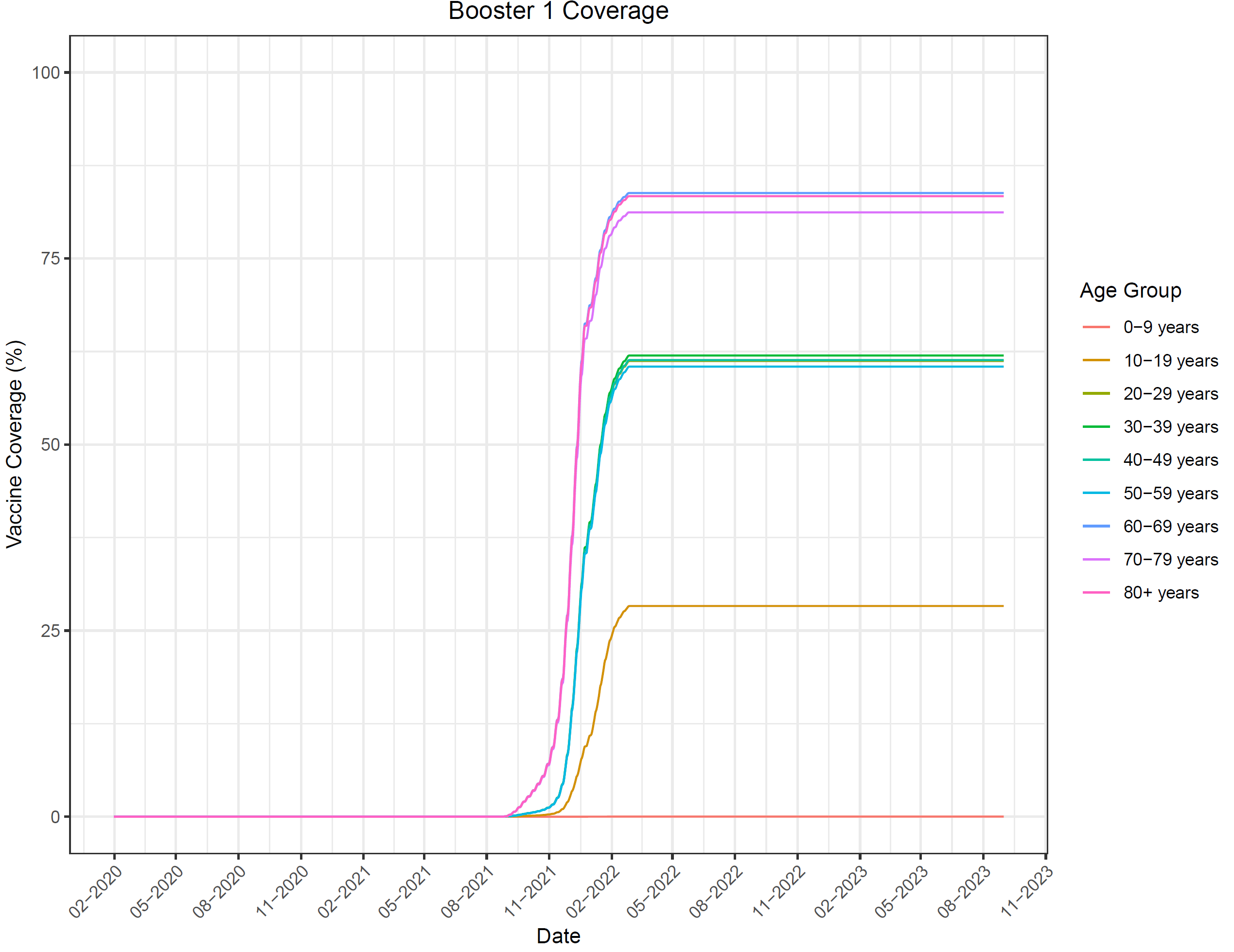

Figure 4. Percentage of the population who received two boosters, by age group

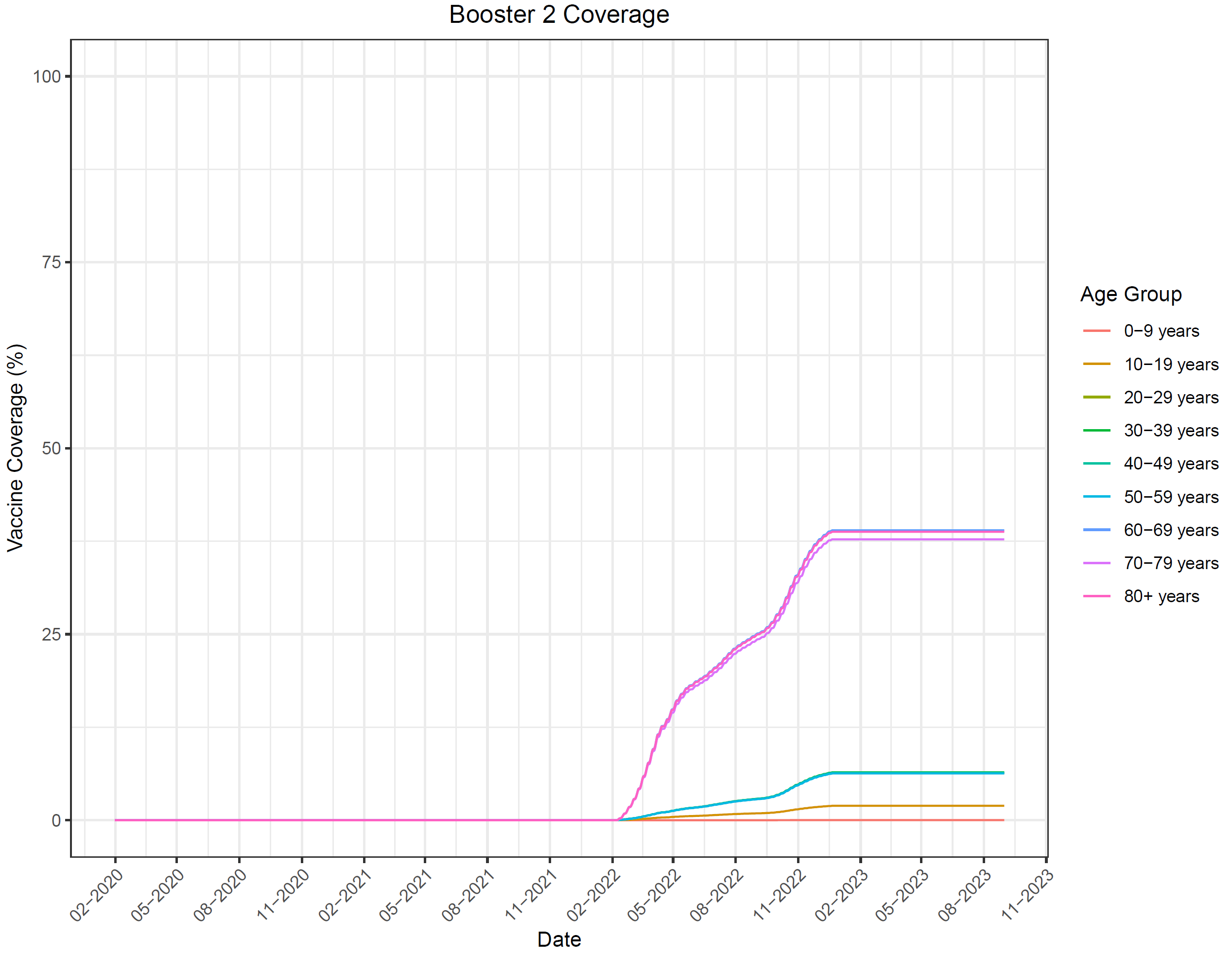

Figure 5. Percentage of the population who received a third booster, by age group

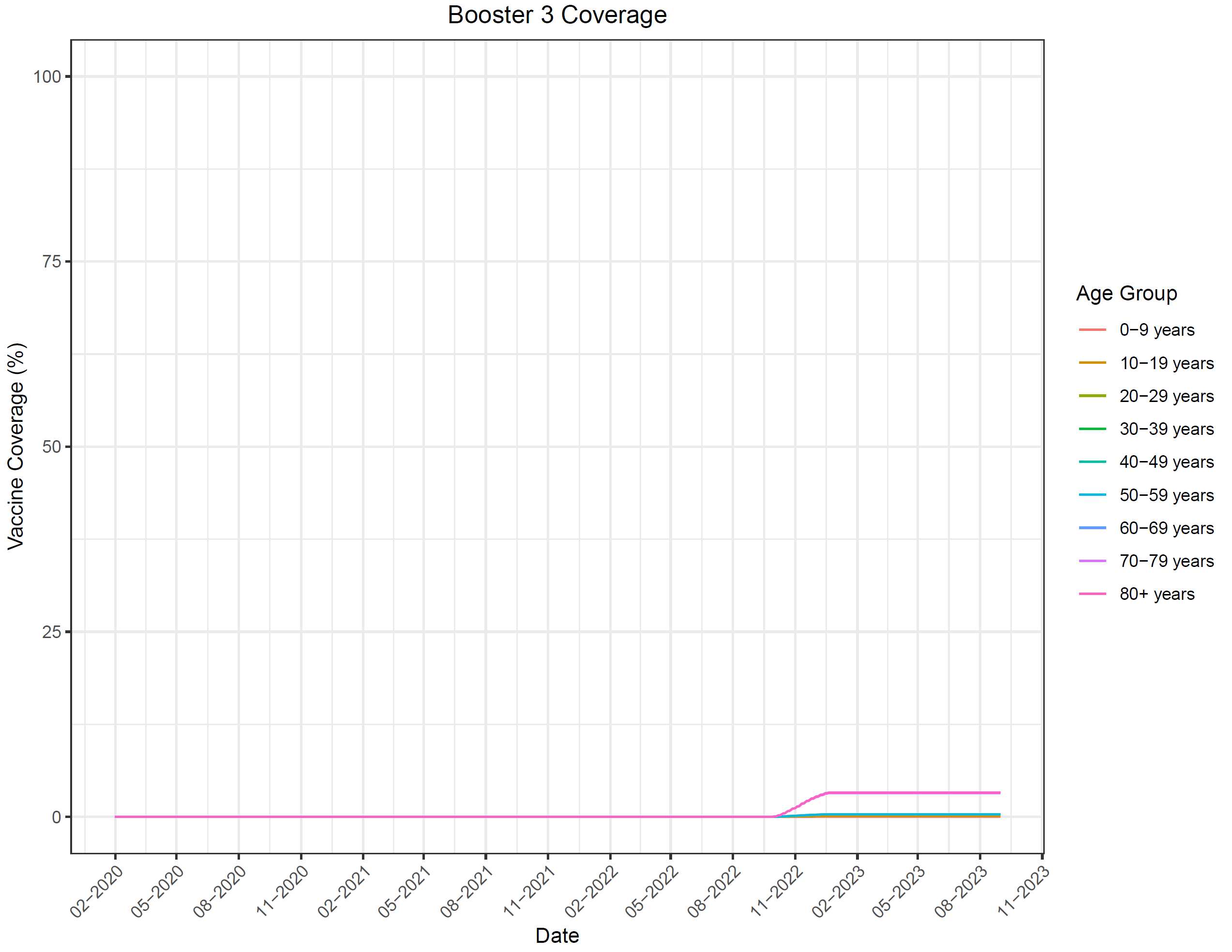

#### 1.4. Other Model Inputs

The average length of time spent by infected individuals exposed to the virus before they become infectious (e.g., latent period) was assumed to be three days [6]. The length of time spent by an individual in an infectious state (e.g., infectious period) was assumed to be seven days [6,7].

In the base case, the rate of natural immunity waning was set to be equal to the rate of waning of primary series vaccine effectiveness (VE) against infection. Therefore, the monthly rate of waning was 3.9% for the pre-Omicron period and 3.3% for the Omicron and Omicron BA.4/5 periods. When vaccine-mediated immunity decreased on the days that the circulating variant changed (start of the Omicron period; start of the Omicron BA.4/5 period), a similar drop in the number of people in the Recovered compartment was also implemented.

#### 1.5. Transmissibility: February 2020 to August 2023

During the first 30 days of the burn-in period, an initial 19,872 infections were used to seed or start the pandemic. This number is equal to the number of infections estimated by the IHME over the first 30 days [4]. Model calibration was then conducted to estimate the transmissibility parameter that reflects cases of COVID-19 experienced in Germany from January 2020--August 2023. The analytical choices for the calibration process were made considering the recommendations of Vanni and colleagues [8]. The transmissibility parameter was allowed to vary on a daily basis. For the calibration process, transmissibility parameters were manually varied. The calibration targets and the goodness of fit measures used, are described together with the results of the calibration below.

##### 1.5.1. Goodness of Fit Measure

An initial model calibration was performed in a qualitative way by visually comparing the daily incidence of COVID cases predicted by the model and the values obtained from the IHME data for the first time period (4 February 2020 through 15 April 2022). For the second time period of the calibration, 15 May 2022 through 15 April 2023, the daily incidence of COVID 19 cases reported by RKI, adjusted to account for asymptomatic infections was compared to the cases predicted by the model. Between 15 April 2022 and 15 May 2022 a linear interpolation between COVID cases predicted by IHME and those reported by RKI was compared to cases predicted by the model.

##### 1.5.2. Results of the Calibration

The final transmissibility parameters for the calibration period for the base case scenario are summarized in Table 3. For the dates shown, the transmissibility parameter was manually varied and linear interpolation was used to estimate values for the days between these dates. The comparison plot of the daily number of incident cases of COVID predicted from the calibrated model and the corresponding estimates from the IHME is presented in Figure 6. The comparison of the monthly estimates of the number of hospitalizations is presented in Figure 7.

Table 3. Final transmissibility parameter inputs for the SEIR model

| **Date** | **Days from start of analysis** | **Transmissibility parameter estimate** |
| --- | --- | --- |
| January 31, 2020 | 1 | 0.30 |
| February 29, 2020 | 30 | 0.32 |
| March 15, 2020 | 45 | 0.35 |
| March 23, 2020 | 53 | 0.41 |
| March 31, 2020 | 61 | 0.24 |
| April 15, 2020 | 76 | 0.16 |
| April 30, 2020 | 91 | 0.15 |
| May 31, 2020 | 122 | 0.15 |
| June 30, 2020 | 152 | 0.18 |
| July 31, 2020 | 183 | 0.16 |
| August 31, 2020 | 214 | 0.20 |
| September 30, 2020 | 244 | 0.24 |
| October 31, 2020 | 275 | 0.25 |
| November 15, 2020 | 290 | 0.27 |
| November 30, 2020 | 305 | 0.27 |
| December 15, 2020 | 320 | 0.27 |
| December 25, 2020 | 330 | 0.38 |
| December 31, 2020 | 336 | 0.35 |
| January 9, 2021 | 345 | 0.35 |
| January 11, 2021 | 347 | 0.40 |
| January 14, 2021 | 350 | 0.28 |
| January 19, 2021 | 355 | 0.20 |
| January 31, 2021 | 367 | 0.20 |
| February 08, 2021 | 375 | 0.23 |
| February 28, 2021 | 395 | 0.26 |
| March 10, 2021 | 405 | 0.29 |
| March 20, 2021 | 415 | 0.29 |
| March 31, 2021 | 426 | 0.26 |
| April 9, 2021 | 435 | 0.25 |
| April 24, 2021 | 450 | 0.22 |
| May 9, 2021 | 465 | 0.16 |
| May 14, 2021 | 470 | 0.16 |
| May 16, 2021 | 472 | 0.15 |
| May 24, 2021 | 480 | 0.13 |
| May 31, 2021 | 487 | 0.13 |
| June 8, 2021 | 495 | 0.10 |
| June 13, 2021 | 500 | 0.12 |
| June 18, 2021 | 505 | 0.12 |
| June 23, 2021 | 510 | 0.30 |
| June 28, 2021 | 515 | 0.25 |
| July 3, 2021 | 520 | 0.25 |
| July 13, 2021 | 530 | 0.25 |
| July 23, 2021 | 540 | 0.25 |
| August 2, 2021 | 550 | 0.30 |
| August 12, 2021 | 560 | 0.35 |
| August 22, 2021 | 570 | 0.38 |
| September 1, 2021 | 580 | 0.22 |
| September 11, 2021 | 590 | 0.20 |
| September 21, 2021 | 600 | 0.25 |
| September 26, 2021 | 605 | 0.30 |
| October 1, 2021 | 610 | 0.35 |
| October 11, 2021 | 620 | 0.39 |
| November 10, 2021 | 650 | 0.40 |
| November 20, 2021 | 660 | 0.27 |
| November 25, 2021 | 665 | 0.27 |
| November 30, 2021 | 670 | 0.22 |
| December 10, 2021 | 680 | 0.24 |
| December 20, 2021 | 690 | 0.30 |
| December 25, 2021 | 695 | 0.38 |
| December 30, 2021 | 700 | 0.38 |
| January 2, 2022 | 703 | 0.42 |
| January 3, 2022 | 704 | 0.40 |
| January 4, 2022 | 705 | 0.50 |
| January 5, 2022 | 706 | 0.52 |
| January 14, 2022 | 715 | 0.48 |
| January 19, 2022 | 720 | 0.46 |
| January 24, 2022 | 725 | 0.45 |
| January 29, 2022 | 730 | 0.40 |
| January 31, 2022 | 732 | 0.40 |
| February 8, 2022 | 740 | 0.40 |
| February 14, 2022 | 746 | 0.38 |
| February 23, 2022 | 755 | 0.42 |
| February 26, 2022 | 758 | 0.48 |
| February 28, 2022 | 760 | 0.48 |
| March 5, 2022 | 765 | 0.5 |
| March 10, 2022 | 770 | 0.57 |
| March 15, 2022 | 775 | 0.57 |
| March 20, 2022 | 780 | 0.58 |
| March 25, 2022 | 785 | 0.52 |
| March 31, 2022 | 791 | 0.65 |
| April 4, 2022 | 795 | 0.65 |
| April 7, 2022 | 798 | 0.67 |
| April 9, 2022 | 800 | 0.73 |
| April 10, 2022 | 801 | 0.77 |
| April 11, 2022 | 802 | 0.77 |
| April 14, 2022 | 805 | 0.85 |
| April 15, 2022 | 806 | 0.85 |
| April 16, 2022 | 807 | 0.85 |
| April 17, 2022 | 808 | 0.85 |
| April 19, 2022 | 810 | 0.85 |
| April 24, 2022 | 815 | 0.80 |
| April 28, 2022 | 819 | 0.80 |
| April 30, 2022 | 821 | 0.70 |
| May 2, 2022 | 823 | 0.65 |
| May 4, 2022 | 825 | 0.65 |
| May 6, 2022 | 827 | 0.65 |
| May 8, 2022 | 829 | 0.65 |
| May 9, 2022 | 830 | 0.65 |
| May 10, 2022 | 831 | 0.12 |
| May 12, 2022 | 833 | 0.10 |
| May 15, 2022 | 836 | 0.09 |
| May 17, 2022 | 838 | 0.09 |
| May 19, 2022 | 840 | 0.09 |
| May 21, 2022 | 842 | 0.09 |
| May 23, 2022 | 844 | 0.10 |
| May 25, 2022 | 846 | 0.10 |
| May 29, 2022 | 850 | 0.17 |
| May 31, 2022 | 852 | 0.23 |
| June 3, 2022 | 855 | 0.28 |
| June 8, 2022 | 860 | 0.32 |
| June 15, 2022 | 867 | 0.34 |
| June 23, 2022 | 875 | 0.33 |
| June 30, 2022 | 882 | 0.31 |
| July 3, 2022 | 885 | 0.31 |
| July 8, 2022 | 890 | 0.29 |
| July 13, 2022 | 895 | 0.28 |
| July 23, 2022 | 905 | 0.24 |
| July 28, 2022 | 910 | 0.23 |
| August 2, 2022 | 915 | 0.21 |
| August 7, 2022 | 920 | 0.21 |
| August 12, 2022 | 925 | 0.20 |
| August 14, 2022 | 927 | 0.20 |
| August 17, 2022 | 930 | 0.20 |
| August 22, 2022 | 935 | 0.18 |
| August 25, 2022 | 938 | 0.18 |
| August 27, 2022 | 940 | 0.20 |
| August 29, 2022 | 942 | 0.20 |
| September 6, 2022 | 950 | 0.24 |
| September 11, 2022 | 955 | 0.28 |
| September 16, 2022 | 960 | 0.30 |
| September 21, 2022 | 965 | 0.32 |
| September 26, 2022 | 970 | 0.34 |
| October 3, 2022 | 977 | 0.39 |
| October 6, 2022 | 980 | 0.39 |
| October 10, 2022 | 984 | 0.33 |
| October 21, 2022 | 995 | 0.22 |
| October 31, 2022 | 1005 | 0.17 |
| November 10, 2022 | 1015 | 0.15 |
| November 15, 2022 | 1020 | 0.18 |
| November 17, 2022 | 1022 | 0.18 |
| November 19, 2022 | 1024 | 0.20 |
| November 23, 2022 | 1028 | 0.22 |
| November 25, 2022 | 1030 | 0.23 |
| November 27, 2022 | 1032 | 0.24 |
| November 30, 2022 | 1035 | 0.25 |
| December 5, 2022 | 1040 | 0.27 |
| December 10, 2022 | 1045 | 0.27 |
| December 15, 2022 | 1050 | 0.28 |
| December 17, 2022 | 1052 | 0.27 |
| December 22, 2022 | 1057 | 0.27 |
| December 25, 2022 | 1060 | 0.20 |
| December 30, 2022 | 1065 | 0.16 |
| January 4, 2023 | 1070 | 0.19 |
| January 9, 2023 | 1075 | 0.16 |
| January 24, 2023 | 1090 | 0.16 |
| January 26, 2023 | 1092 | 0.24 |
| January 31, 2023 | 1097 | 0.24 |
| February 8, 2023 | 1105 | 0.26 |
| February 15, 2023 | 1112 | 0.27 |
| February 18, 2023 | 1115 | 0.28 |
| February 23, 2023 | 1120 | 0.28 |
| February 28, 2023 | 1125 | 0.21 |
| March 5, 2023 | 1130 | 0.16 |
| March 10, 2023 | 1135 | 0.11 |
| March 17, 2023 | 1142 | 0.15 |
| March 20, 2023 | 1145 | 0.17 |
| March 25, 2023 | 1150 | 0.19 |
| March 31, 2023 | 1156 | 0.19 |
| April 15, 2023 | 1171 | 0.26 |
| **Start projection period** | | |
| June 2, 2023 | 1217 | 0.20 |
| August 2, 203 | 1278 | 0.23 |
| September 2, 2023 | 1309 | 0.30 |
| December 2, 2023 | 1400 | 0.23 |
| February 2, 2023 | 1462 | 0.20 |
| March 2, 2023 | 1491 | 0.22 |
| August 2, 2023 | 1644 | 0.26 |

Figure 6. Comparison of daily incidence of COVID (calibrated model estimates vs IHME estimates February 2020 to 15 April 2022); RKI estimates adjusted for asymptomatic infections 15 May 2022 to 15 April 2023)

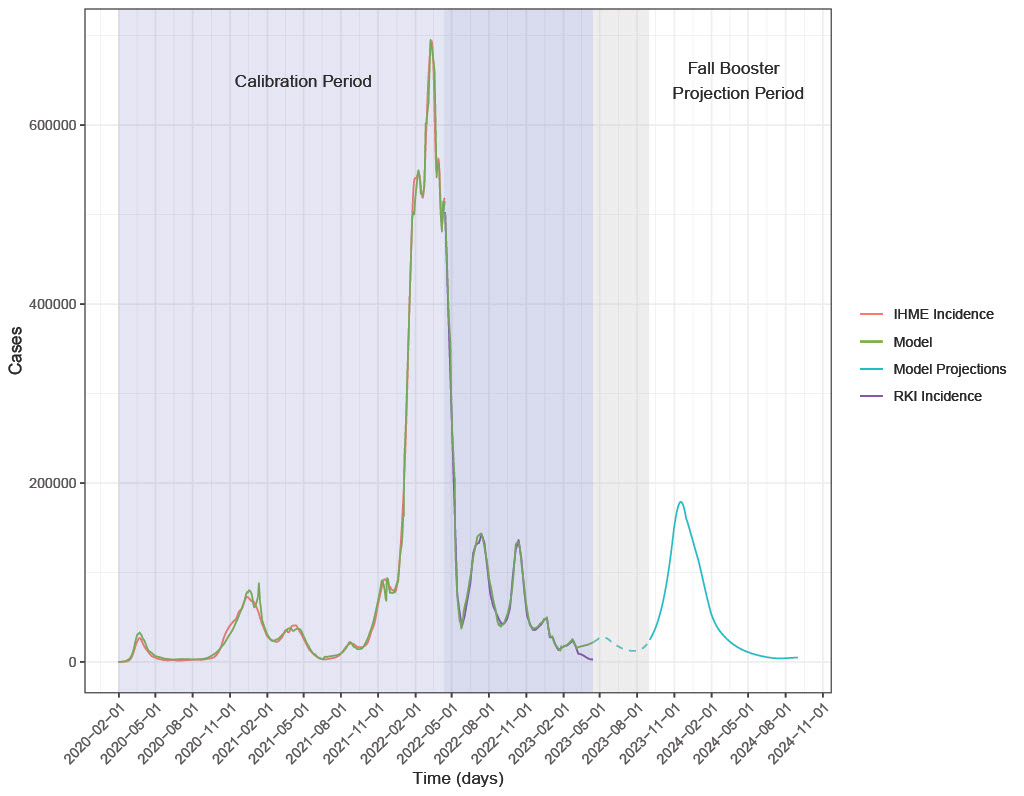

IHME: Institute for Health Metrics and Evaluation; RKI: Robert-Koch Institut

### 2. SEIR Model Inputs: Analytic Time Period (September 2023 to August 2024)

The inputs that were changed for the analytic time horizon are described below. Inputs not described were the same as during the burn-in period.

#### 2.1. Vaccine Coverage

Individuals under 30 years of age are modelled, but their vaccination status does not change after 1 September 2023, and they are not eligible to receive an Autumn booster as this analysis focuses on at-risk adults aged 30-59 years as well as persons aged 60 years and older.

Base case vaccine uptake was based on influenza vaccine uptake for the 2021--2022 season and assumed to occur between September 2023--December 2023 (Figure 7). For the comparison of the mRNA-1273.815 vaccine strategy to no vaccination, a scenario analyses were also included where the vaccine coverage is half of the base case values (Figure 8).

Figure 7. Vaccine coverage, base case

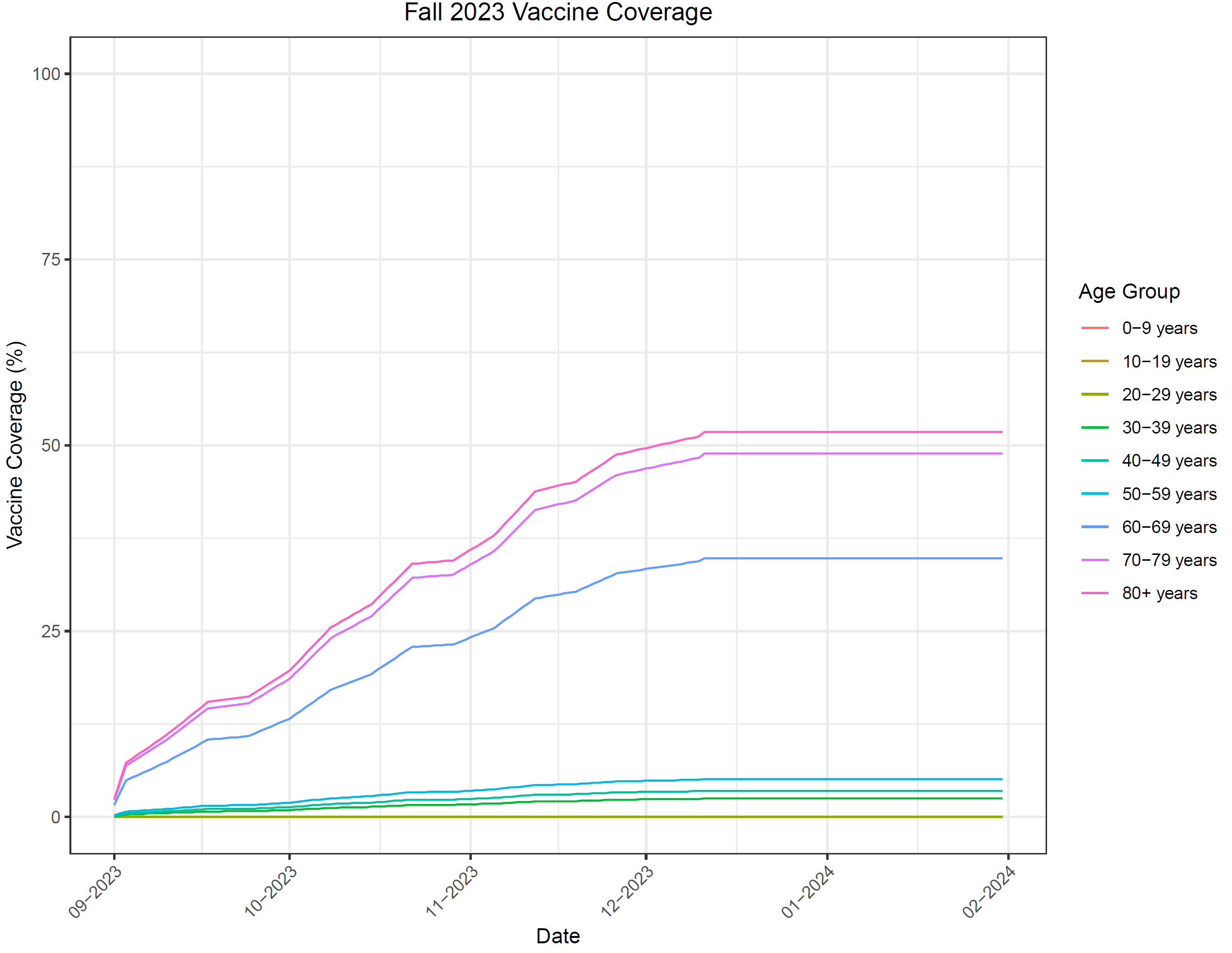

Figure 8. Vaccine coverage, scenario analysis: half of base case values

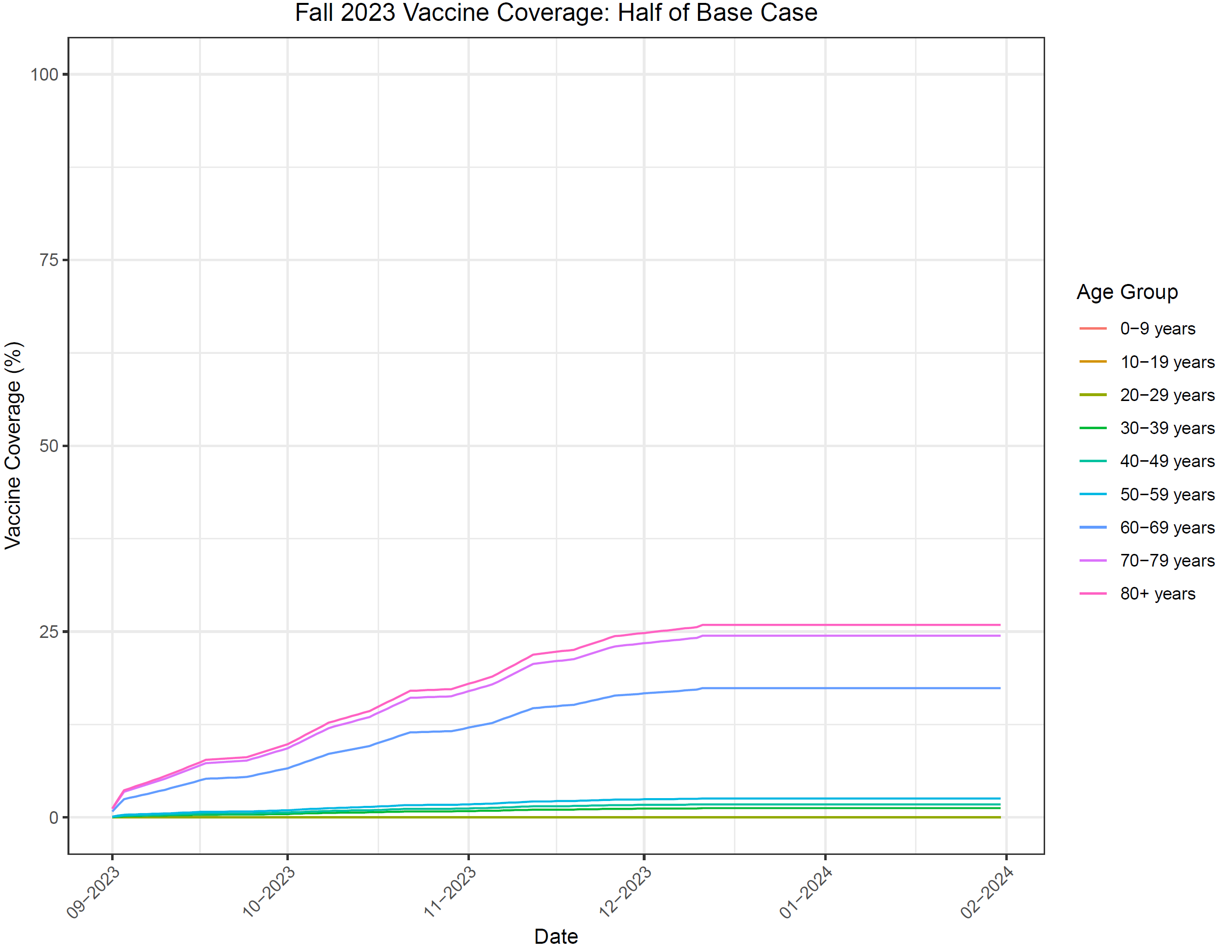

Table 4. Economic analysis results (mRNA-1273 vs BNT162b2)

|  | **Healthcare Costs** | | | **Lost Productivity Costs** | | | **Total Costs** | | |
| --- | --- | --- | --- | --- | --- | --- | --- | --- | --- |
| **Variable** | **BNT162b2** | **mRNA-1273** | **Difference*** | **BNT162b2** | **mRNA-1273** | **Difference*** | **BNT162b2** | **mRNA-1273** | **Difference*** |
| Vaccination | €1,395,681,758 | €1,395,681,758 | €0 | €65,167,523 | €65,167,523 | €0 | €1,460,849,281 | €1,460,849,281 | €0 |
| Short-Term Infection Costs | €3,742,645,933 | €3,683,873,064 | -€58,772,869 | €3,283,469,698 | €3,260,661,141 | -€22,808,557 | €7,026,115,631 | €6,944,534,205 | -€81,581,426 |
| Not Hospitalized | €213,980,900 | €212,310,317 | -€1,670,583 | €3,107,516,833 | €3,086,556,974 | -€20,959,859 | €3,321,497,733 | €3,298,867,291 | -€22,630,442 |
| Hospitalized | | | | | | | | | |
| No ICU or ventilator | €1,049,969,949 | €1,034,245,190 | -€15,724,758 | €94,867,276 | €93,957,831 | -€909,445 | €1,144,837,225 | €1,128,203,222 | -€16,634,203 |
| Initial hospitalization | €1,049,969,949 | €1,034,245,190 | -€15,724,758 |  |  |  | €1,049,969,949 | €1,034,245,190 | -€15,724,758 |
| ICU only | €745,398,326 | €733,233,024 | -€12,165,302 | €18,740,649 | €18,540,022 | -€200,627 | €764,138,975 | €751,773,046 | -€12,365,929 |
| Initial hospitalization | €745,398,326 | €733,233,024 | -€12,165,302 |  |  |  | €745,398,326 | €733,233,024 | -€12,165,302 |
| Ventilator | €1,729,994,837 | €1,700,833,155 | -€29,161,682 | €62,344,941 | €61,606,314 | -€738,627 | €1,792,339,778 | €1,762,439,469 | -€29,900,309 |
| Initial hospitalization | €1,729,994,837 | €1,700,833,155 | -€29,161,682 |  |  |  | €1,729,994,837 | €1,700,833,155 | -€29,161,682 |
| Hospitalization Recovery | €3,301,921 | €3,251,378 | -€50,543 | €0 | €0 | €0 | €3,301,921 | €3,251,378 | -€50,543 |
| **Total** | **€5,138,327,691** | **€5,079,5254,822** | **-€58,772,869** | **€3,348,637,222** | **€3,325,828,664** | **-€22,808,557** | **€8,486,964,913** | **€8,405,383,486** | **-€81,581,426** |

Table 5. Deterministic sensitivity analysis results (mRNA-1273.815 campaign vs no vaccination), healthcare payer perspective

| **Model Parameter** | **Variation** | **ICER**  **(Cost/QALY Gained)** |
| --- | --- | --- |
| Incidence | Double the waning rate for natural immunity during Omicron period | mRNA-1273.815 dominates |
| Incidence | Half the waning rate for natural immunity during Omicron period | €904 |
| Vaccine coverage | Uptake half of base-case | mRNA-1273.815 dominates |
| mRNA-1273.815 vaccine effectiveness | Decreased initial VE (Lower 95% CI) against infection | €5,375 |
| mRNA-1273.815 vaccine effectiveness | Increased initial VE (Upper 95% CI) against infection | mRNA-1273.815 dominates |
| mRNA-1273.815 vaccine effectiveness | Decreased initial VE (Lower 95% CI) against hospitalization | €243 |
| mRNA-1273.815 vaccine effectiveness | Increased initial VE (Upper 95% CI) against hospitalization | mRNA-1273.815 dominates |
| mRNA-1273.815 vaccine effectiveness | Decreased vaccine waning (Outcome: Infection) | mRNA-1273.815 dominates |
| mRNA-1273.815 vaccine effectiveness | Increased vaccine waning (Outcome: Infection) | €375 |
| mRNA-1273.815 vaccine effectiveness | Decreased vaccine waning (Outcome: hospitalization) | mRNA-1273.815 dominates |
| mRNA-1273.815 vaccine effectiveness | Increased vaccine waning (Outcome: hospitalization) | €12 |
| mRNA-1273.815 vaccine effectiveness | Decreased vaccine waning | mRNA-1273.815 dominates |
| mRNA-1273.815 vaccine effectiveness | Increased vaccine waning | €904 |

CI: confidence interval; ICER: incremental cost-effectiveness ratio; QALY: quality-adjusted life-year; VE: vaccine effectiveness

Figure 9. Deterministic sensitivity analyses from payer and societal perspectives

Tornado plot for Return on Investment mRNA 1273.851 vs. No Vaccination (Payer Perspective)

Return on Investment

| Varies Parameter (Sets) in Sensitivity Analysis | Upper Values | Lower Values |
| --- | --- | --- |
| Mortality Rate | 1.04087 | 1.04088 |
| Outpatient Rate | 1.04533 | 1.03642 |
| Proportion Symptomatic | 1.04557 | 1.03618 |
| Outpatient Price | 1.04682 | 1.03493 |
| Waning VE against Hospitalization | 1.00642 | 1.0683 |
| Waning VE against Infection | 1.00642 | 1.09022 |
| VE against Hospitalization | 1.08113 | 0.990554 |
| Hospitalization Rate | 1.15844 | 0.923305 |
| mRNA 1273.851 Price | 0.879716 | 1.27432 |
| Hospitalization Price | 1.24292 | 0.838826 |
| VE against Infection | 1.27485 | 0.804613 |

Tornado plot for Benefit-Cost-Ratio (BCR) mRNA 1273.851 vs. No Vaccination (Payer Perspective)

Benefit-Cost-Ratio (BCR)

| Varies Parameter (Sets) in Sensitivity Analysis | Upper Values | Lower Values |
| --- | --- | --- |
| Outpatient Rate | 4.51808 | 4.50917 |
| Outpatient Price | 4.51957 | 4.50768 |
| Waning VE against Hospitalization | 4.43524 | 4.57601 |
| VE against Hospitalization | 4.60516 | 4.39921 |
| Hospitalization Rate | 4.63398 | 4.39327 |
| Mortality Rate | 4.70248 | 4.32477 |
| Hospitalization Price | 4.71567 | 4.31158 |
| Waning VE against Infection | 4.43524 | 4.88825 |
| Proportion Symptomatic | 4.95726 | 4.06999 |
| Utilities | 5.20818 | 3.81908 |
| mRNA 1273.851 Price | 3.81478 | 5.52594 |
| VE against Infection | 6.30937 | 2.67601 |

VE: vaccine effectiveness

Tornado plot for Return on Investment mRNA 1273.851 vs. No Vaccination (Societal Perspective)

Return on Investment

| Varies Parameter (Sets) in Sensitivity Analysis | Upper Values | Lower Values |
| --- | --- | --- |
| Mortality Rate | 1.38691 | 1.38572 |
| Outpatient Price | 1.39204 | 1.38059 |
| Outpatient Rate | 1.40039 | 1.37224 |
| Waning VE against Hospitalization | 1.35273 | 1.41305 |
| VE against Hospitalization | 1.42555 | 1.33727 |
| Waning VE against Infection | 1.35273 | 1.48386 |
| Productivity Loss Days | 1.45168 | 1.31995 |
| Proportion Symptomatic | 1.46394 | 1.3087 |
| Hospitalization Rate | 1.50046 | 1.27217 |
| Productivity Loss Price | 1.53099 | 1.26463 |
| Hospitalization Price | 1.58069 | 1.19194 |
| mRNA 1273.851 Price | 1.17861 | 1.6829 |
| VE against Infection | 1.84964 | 0.924193 |

Tornado plot for Benefit-Cost-Ratio (BCR) mRNA 1273.851 vs. No Vaccination (Societal Perspective)

Benefit-Cost-Ratio (BCR)

| Varies Parameter (Sets) in Sensitivity Analysis | Upper Values | Lower Values |
| --- | --- | --- |
| OutpatientPrice | 4.73287 | 4.72142 |
| OutpatientRate | 4.74122 | 4.71307 |
| ProductivityLossDays | 4.76732 | 4.68636 |
| WaningVEhosp | 4.65131 | 4.78751 |
| ProductivityLossPrice | 4.8169 | 4.65178 |
| VEhosp | 4.81571 | 4.61644 |
| HospitalizationRate | 4.84397 | 4.61032 |
| MortalityRate | 4.90943 | 4.54486 |
| HospitalizationPrice | 4.92152 | 4.53277 |
| WaningVEinf | 4.65131 | 5.13761 |
| Symptomatic | 5.22703 | 4.22726 |
| Utilities | 5.39531 | 4.05898 |
| mRNA 1273.851 Price | 4.01888 | 5.73846 |
| VEinf | 6.69291 | 2.7245 |

VE: vaccine effectiveness

### 3. Equations associated with the SEIR Model

#### 3.1. Differential Equations

The following equations describe the movement of people in the population through the SEIR compartments and the vaccination strata.

Unvaccinated cohort

$$S_{t+1,j}^{X}=S_{t,j}^{X}-\lambda_{t,j}^{X}S_{t,j}^{X}-\left( p_{t,j}^{X,S} \right)\mu_{t,j}+\omega_{t,j}^{X}R_{t,j}^{X}-\epsilon_{t,j}$$

$$E_{t+1,j}^{X}=E_{t,j}^{X}+\lambda_{t,j}^{X}S_{t,j}^{X}-\frac{1}{\tau_{E}}E_{t,j}^{X}$$

$$I_{t+1,j}^{X}=I_{t,j}^{X}+\frac{1}{\tau_{E}}E_{t,j}^{X}-\frac{1}{\tau_{I}}I_{t,j}^{X}+\epsilon_{t,j}$$

$$R_{t+1,j}^{X}=R_{t,j}^{X}-\left( p_{t,j}^{X,R} \right)\mu_{t,j}+\frac{1}{\tau_{I}}I_{t,j}^{X}-\omega_{t,j}^{X}R_{t,j}^{X}$$

Vaccinated cohort

$$S_{t+1,j}^{V}=S_{t,j}^{V}-\lambda_{t,j}^{V}S_{t,j}^{V}+{\left( p_{t,j}^{X,S} \right)\mu}_{t,j}-\left( p_{t,j}^{V,S} \right)\nu_{t,j}-\left( p_{t,j}^{V,S} \right)p_{t,j}^{V,B3}\nu_{t,j}^{3}-\left( p_{t,j}^{V,S} \right)p_{t,j}^{V,B4}\nu_{t,j}^{4}+\omega_{t,j}^{V}R_{t,j}^{V}$$

$$E_{t+1,j}^{V}=E_{t,j}^{V}+\lambda_{t,j}^{V}S_{t,j}^{V}-\frac{1}{\tau_{E}}E_{t,j}^{V}$$

$$I_{t+1,j}^{V}=I_{t,j}^{V}+\frac{1}{\tau_{E}}E_{t,j}^{V}-\frac{1}{\tau_{I}}I_{t,j}^{V}$$

$$R_{t+1,j}^{V}=R_{t,j}^{V}+{\left( p_{t,j}^{X,R} \right)\mu}_{t,j}-\left( p_{t,j}^{V,R} \right)\nu_{t,j}-\left( p_{t,j}^{V,R} \right)p_{t,j}^{V,B3}\nu_{t,j}^{3}-\left( p_{t,j}^{V,R} \right)p_{t,j}^{V,B4}\nu_{t,j}^{4}+\frac{1}{\tau_{I}}I_{t,j}^{V}-\omega_{t,j}^{V}R_{t,j}^{V}$$

First Booster cohort

$$S_{t+1,j}^{B}=S_{t,j}^{B}-\lambda_{t,j}^{B}S_{t,j}^{B}+\left( p_{t,j}^{V,S} \right)\nu_{t,j}-{\left( p_{t,j}^{B,S} \right)\nu}_{t,j}^{2}-{\left( p_{t,j}^{B,S} \right)\left( p_{t,j}^{B,B3} \right)\nu}_{t,j}^{3}-{\left( p_{t,j}^{B,S} \right)\left( p_{t,j}^{B,B4} \right)\nu}_{t,j}^{4}+\omega_{t,j}^{B}R_{t,j}^{B}$$

$$E_{t+1,j}^{B}=E_{t,j}^{B}+\lambda_{t,j}^{B}S_{t,j}^{B}-\frac{1}{\tau_{E}}E_{t,j}^{B}$$

$$I_{t+1,j}^{B}=I_{t,j}^{B}+\frac{1}{\tau_{E}}E_{t,j}^{B}-\frac{1}{\tau_{I}}I_{t,j}^{B}$$

$$R_{t+1,j}^{B}=R_{t,j}^{B}+\left( p_{t,j}^{V,R} \right)\nu_{t,j}-{\left( p_{t,j}^{B,R} \right)\nu}_{t,j}^{2}-{\left( p_{t,j}^{B,R} \right)\left( p_{t,j}^{B,B3} \right)\nu}_{t,j}^{3}-{\left( p_{t,j}^{B,R} \right)\left( p_{t,j}^{B,B4} \right)\nu}_{t,j}^{4}+\frac{1}{\tau_{I}}I_{t,j}^{B}-\omega_{t,j}^{B}R_{t,j}^{B}$$

Second Booster cohort

$$S_{t+1,j}^{B2}=S_{t,j}^{B2}-\lambda_{t,j}^{B2}S_{t,j}^{B2}+{\left( p_{t,j}^{B,S} \right)\nu}_{t,j}^{2}-\left( p_{t,j}^{B2,S} \right)\left( p_{t,j}^{B2,B3} \right)\nu_{t,j}^{3}-\left( p_{t,j}^{B2,S} \right)\left( p_{t,j}^{B2,B4} \right)\nu_{t,j}^{4}+\omega_{t,j}^{B2}R_{t,j}^{B2}$$

$$E_{t+1,j}^{B2}=E_{t,j}^{B2}+\lambda_{t,j}^{B2}S_{t,j}^{B2}-\frac{1}{\tau_{E}}E_{t,j}^{B2}$$

$$I_{t+1,j}^{B2}=I_{t,j}^{B2}+\frac{1}{\tau_{E}}E_{t,j}^{B2}-\frac{1}{\tau_{I}}I_{t,j}^{B2}$$

$$R_{t+1,j}^{B2}=R_{t,j}^{B2}+{\left( p_{t,j}^{B,R} \right)\nu}_{t,j}^{2}-\left( p_{t,j}^{B2,R} \right)\left( p_{t,j}^{B2,B3} \right)\nu_{t,j}^{3}-\left( p_{t,j}^{B2,R} \right)\left( p_{t,j}^{B2,B4} \right)\nu_{t,j}^{4}+\frac{1}{\tau_{I}}I_{t,j}^{B2}-\omega_{t,j}^{B2}R_{t,j}^{B2}$$

Third Booster cohort

$$S_{t+1,j}^{B3}=S_{t,j}^{B3}-\lambda_{t,j}^{B3}S_{t,j}^{B3}+\left( \left( p_{t,j}^{V,S} \right)p_{t,j}^{V,B3}+\left( p_{t,j}^{B,S} \right)p_{t,j}^{B,B3}+{\left( p_{t,j}^{B2,S} \right)p}_{t,j}^{B2,B3} \right)\nu_{t,j}^{3}-\left( p_{t,j}^{B3,S} \right)\left( p_{t,j}^{B3,B4} \right)\nu_{t,j}^{4}+\omega_{t,j}^{B3}R_{t,j}^{B3}$$

$$E_{t+1,j}^{B3}=E_{t,j}^{B3}+\lambda_{t,j}^{B3}S_{t,j}^{B3}-\frac{1}{\tau_{E}}E_{t,j}^{B3}$$

$$I_{t+1,j}^{B3}=I_{t,j}^{B3}+\frac{1}{\tau_{E}}E_{t,j}^{B3}-\frac{1}{\tau_{I}}I_{t,j}^{B3}$$

$$R_{t+1,j}^{B3}=R_{t,j}^{B3}+\left( \left( p_{t,j}^{V,R} \right)p_{t,j}^{V,B3}+\left( p_{t,j}^{B,R} \right)p_{t,j}^{B,B3}+{\left( p_{t,j}^{B2,R} \right)p}_{t,j}^{B2,B3} \right)\nu_{t,j}^{3}-\left( p_{t,j}^{B3,R} \right)\left( p_{t,j}^{B3,B4} \right)\nu_{t,j}^{4}+\frac{1}{\tau_{I}}I_{t,j}^{B3}-\omega_{t,j}^{B3}R_{t,j}^{B3}$$

Fourth Boosted cohort

$$S_{t+1,j}^{B4}=S_{t,j}^{B4}-\lambda_{t,j}^{B4}S_{t,j}^{B4}+\left( \left( p_{t,j}^{V,S} \right)p_{t,j}^{V,B4}+\left( p_{t,j}^{B,S} \right)p_{t,j}^{B,B4}+{\left( p_{t,j}^{B2,S} \right)p}_{t,j}^{B2,B4}+{\left( p_{t,j}^{B3,S} \right)p}_{t,j}^{B3,B4} \right)\nu_{t,j}^{4}+\omega_{t,j}^{B4}R_{t,j}^{B4}$$

$$E_{t+1,j}^{B4}=E_{t,j}^{B4}+\lambda_{t,j}^{B4}S_{t,j}^{B4}-\frac{1}{\tau_{E}}E_{t,j}^{B4}$$

$$I_{t+1,j}^{B4}=I_{t,j}^{B4}+\frac{1}{\tau_{E}}E_{t,j}^{B4}-\frac{1}{\tau_{I}}I_{t,j}^{B4}$$

$$R_{t+1,j}^{B4}=R_{t,j}^{B4}+\left( \left( p_{t,j}^{V,R} \right)p_{t,j}^{V,B4}+\left( p_{t,j}^{B,R} \right)p_{t,j}^{B,B4}+{\left( p_{t,j}^{B2,R} \right)p}_{t,j}^{B2,B4}+{\left( p_{t,j}^{B3,R} \right)p}_{t,j}^{B3,B4} \right)\nu_{t,j}^{4}+\frac{1}{\tau_{I}}I_{t,j}^{B4}-\omega_{t,j}^{B4}R_{t,j}^{B4}$$

For t=1,

$S_{1,j}^{X}$ = Initial number of susceptible individuals

$I_{1,j}^{X}$= Initial number of infectious individuals = $\epsilon_{1,j}$

$$E_{1,j}^{X}=R_{1,j}^{X}=0$$

$$S_{1,j}^{V}=E_{1,j}^{V}=I_{1,j}^{V}=R_{1,j}^{V}=0$$

$$S_{1,j}^{B}=E_{1,j}^{B}=I_{1,j}^{B}=R_{1,j}^{B}=0$$

$$S_{1,j}^{B2}=E_{1,j}^{B2}=I_{1,j}^{B2}=R_{1,j}^{B2}=0$$

$$S_{1,j}^{B3}=E_{1,j}^{B3}=I_{1,j}^{B3}=R_{1,j}^{B3}=0$$

$$S_{1,j}^{B4}=E_{1,j}^{B4}=I_{1,j}^{B4}=R_{1,j}^{B4}=0$$

Notes, superscripts:

*X*, *V*, *B*, *B2*, *B3*, and *B4* represent the unvaccinated, vaccinated, first booster, second booster, third booster (bivalent), and fourth booster (fall 2023 bivalent) cohorts, respectively.

Notes, subscripts:

*t* = time (i.e., day of analysis)

*j* = age group (number 1 to 9)

Definitions

$S_{j}^{X}, S_{j}^{V}, S_{j}^{B}, S_{j}^{B2},S_{j}^{B3},S_{j}^{B4}$ represent the proportion of susceptibles in age group j in cohort X, V, B, B2, B3, or B4.

The compartments with superscript *X*, *V*, *B, B1, B2, B3, and B4* represent the unvaccinated, vaccinated, and boosted cohorts, respectively.

$E_{j}^{Z}$ represent exposed, but not yet infectious, individuals in age group *j* in cohort *Z* (*X*, *V*, *B,* *B2*, *B3*, or *B4*).

$I_{j}^{Z}$ represent infectious individuals in age group *j* in cohort *Z* (*X*, *V*, *B,* *B2*, *B3*, or *B4*).

$R_{j}^{Z}$ represent immune individuals in age group *j* in cohort *Z* (*X*, *V*, *B,* *B2*, *B3*, or *B4*).

$\lambda_{t,j}^{*}$is the age-group specific force of infection (see section below)

$\frac{1}{\tau_{E}}$ is the rate of loss of latency

$\frac{1}{\tau_{I}}$ is the rate of loss of infectiousness

$\mu_{t,j}$ is the proportion receiving a (primary series) vaccination on day *t* in age group *j*

$\nu_{t,j}$ is the proportion receiving a booster on day *t* in age group *j*

$\nu_{t,j}^{2}$ is the proportion receiving a second booster on day *t* in age group *j*

$\nu_{t,j}^{3}$ is the proportion receiving a third booster on day *t* in age group *j*

$\nu_{t,j}^{4}$ is the proportion receiving a fourth booster on day *t* in age group *j*

$p_{t,j}^{V,B3}$ is the proportion receiving a third booster from cohort *V* on day *t* in age group *j*

$p_{t,j}^{B,B3}$ is the proportion receiving a third booster from cohort *B* on day *t* in age group *j*

$p_{t,j}^{B2,B3}$ is the proportion receiving a third booster from cohort *B2* on day *t* in age group *j*

$p_{t,j}^{V,B4}$ is the proportion receiving a fourth booster from cohort *V* on day *t* in age group *j*

$p_{t,j}^{B,B4}$ is the proportion receiving a fourth booster from cohort *B* on day *t* in age group *j*

$p_{t,j}^{B2,B4}$ is the proportion receiving a fourth booster from cohort *B2* on day *t* in age group *j*

$p_{t,j}^{B3,B4}$ is the proportion receiving a fourth booster from cohort *B3* on day *t* in age group *j*

Note: $p_{t,j}^{V,B3}+p_{t,j}^{B,B3}+p_{t,j}^{B2,B3}=1$

Note: $p_{t,j}^{V,B4}+p_{t,j}^{B,B4}+p_{t,j}^{B2,B4}+p_{t,j}^{B3,B4}=1$

$p_{t-1,j}^{Z,S}$ is the proportion of the booster cohort *Z in the S* compartment out of the total proportion of the booster cohort *Z in the S and R* compartments on day *t-1* in age group *j*

$p_{t-1,j}^{Z,R}$ is the proportion of the booster cohort *Z in the R* compartment out of the total proportion of the booster cohort *Z in the S and R* compartments on day *t-1* in age group *j*

Note: $p_{t-1,j}^{Z,S}+p_{t-1,j}^{Z,R}=1$

$\omega_{t,j}^{Z}$ is the natural immunity waning rate on day *t* in age group *j* in cohort Z (*X*, *V*, *B,* *B2*, *B3*, or *B4)*

$\epsilon_{t,j}$ is the proportion of external cases on day *t* in age group *j*

#### 3.2. Force of infection (unvaccinated)

$$\lambda_{t,i}^{*}=\left( {Overall Scaling Factor}_{t} \right)\times\left[ \beta_{t}\sum_{j=1}^{9} \sum_{Z=\left\{ X,V,B \right\}} c_{ij}I_{t,j}^{Z} \right]$$

$\lambda_{t,i}^{*}$is the age-group specific force of infection at time *t* for age group *i*

${Overall Scaling Factor}_{t}$at time *t* is defined in section 1.2.2.

$\beta_{t}$ is the transmissibility parameter at time *t*

$c_{ij}$ is the rate at which individuals in age group *i* make contact with those in age group *j*

$I_{t,j}^{Z}$ represents the infectious individuals in cohort *Z* (*X*, *V*, *B,* *B2*, *B3*, or *B4*).at time *t* for age group *j*.

#### 3.3. Force of infection (vaccinated)

For the vaccinated cohorts, the force of infection calculation is adjusted based on the VE in the cohort:

$$\lambda_{t,i}^{Z}=\left( 1-{VE}_{t,i}^{X} \right)\times\left( {Overall Scaling Factor}_{t} \right)\times\left[ \beta_{t}\sum_{j=1}^{9} \sum_{Z=\left\{ X,V,B \right\}} c_{ij}I_{t,j}^{Z} \right]$$

$\lambda_{t,i}^{Z}$is the age-group specific force of infection at time *t* for age group *i* in cohort *Z* (*X*, *V*, *B,* *B2*, *B3*, or *B4*).

${VE}_{t,i}^{Z}$ is the vaccine effectiveness at time *t* for age group *i* in cohort *Z* (*X*, *V*, *B,* *B2*, *B3*, or *B4*).The vaccine effectiveness at time *t* is defined in the next section.

#### 3.4. Daily vaccine effectiveness calculations

If no one is vaccinated in the cohort and age group, a VE value of zero is assumed (i.e., when day *t* is less than the day of the beginning of the vaccination period). Once people have been vaccinated in the vaccination cohort and age group, the average vaccine effectiveness on day *t* is calculated as:

$${VE}_{t,i}^{Z}=\frac{\left[ \begin{aligned} \left( Number newly vaccinated on day t\times initial VE \right)+ \\ \left( \left( {VE Drop}_{t} \right)\left( Number previously vaccinated on day t\times({VE}_{t-1,i}^{Z}-daily waning rate \right) \right) \end{aligned} \right]}{\left[ Total number in cohort Z on day t \right]}$$

${VE}_{t,i}^{Z}$ is the vaccine effectiveness at time *t* for age group *i* in cohort *Z* (*X*, *V*, *B,* *B2*, *B3* or *B4*).

If the term $\left( {VE}_{t-1,i}^{Z}-daily waning rate \right)$ falls below zero, we assume a value of zero instead.

The term $\left( {VE Drop}_{t} \right)$ represents a drop in the vaccine effectiveness when a new strain with immune escape enters the population. There are only a few days in which this drop occurs over the course of the time horizon: if a new variant with immune escape emerges, the impact is assumed to happen on the individual days. Apart from those days, the value of the term $\left( {VE Drop}_{t} \right)$ is assumed to be one, corresponding to no impact on the average VE calculation.

#### 3.5. Calculation of incremental effectiveness against hospitalization

For each vaccination cohort, age group, and day, we defined the following vaccine effectiveness variables and relationship between the variables. The superscripts and subscripts for vaccination cohort, age group, and day are removed for clarity.

**Definitions**

${VE}_{1}$ = Vaccine effectiveness against infection

${VE}_{2}$ = ‘Total’ Vaccine effectiveness against hospitalization

${VE}_{2}^{*}$= ‘Additional’ Vaccine effectiveness against hospitalization

We assume ${VE}_{2}^{*}=0$ if there is no additional benefit against hospitalization

Define

$$\left[ 1-{VE}_{2} \right]= \left[ 1-{VE}_{1} \right]\times\left[ 1-{VE}_{2}^{*} \right]$$

Isolate and solve for ${VE}_{2}^{*}$

$$\left[ 1-{VE}_{2}^{*} \right]=\frac{\left[ 1-{VE}_{2} \right]}{\left[ 1-{VE}_{1} \right]}$$

$${VE}_{2}^{*}= 1- \frac{\left[ 1-{VE}_{2} \right]}{\left[ 1-{VE}_{1} \right]}$$

**Probabilities in an unvaccinated cohort**

Probability of COVID-19 infection in an unvaccinated cohort

$$p\left( COVID|UnVac \right)$$

Probability that a COVID-19 infection requires hospitalization in an unvaccinated cohort

$$p\left( Hosp|COVID,UnVac \right)$$

Proportion of an unvaccinated cohort with a COVID-19 infection that requires hospitalization

$$p\left( Hosp,COVID|UnVac \right)= p\left( Hosp|COVID,UnVac \right) \times p\left( COVID|UnVac \right)$$

**Probabilities in a vaccinated cohort**

Probability of COVID-19 infection in a vaccinated cohort

$$p\left( COVID|Vac \right)= \left[ 1-{VE}_{1} \right]\times p\left( COVID|UnVac \right)$$

Probability that an COVD-19 infection requires hospitalization in a vaccinated cohort

$$p\left( Hosp|COVID,Vac \right)= \left[ 1-{VE}_{2}^{*} \right]\times p\left( Hosp|COVID,UnVac \right)$$

Proportion of a vaccinated cohort with an COVID infection that requires hospitalization

$$p\left( Hosp,COVID|Vac \right)= p\left( Hosp|COVID,Vac \right) \times p\left( COVID|Vac \right)$$

### 4. References

[1]	Kohli M, Maschio M, Joshi K, et al. The potential clinical impact and cost-effectiveness of the updated COVID-19 mRNA Fall 2023 vaccines in the United States. medRxiv. 2023. DOI 10.1101/2023.09.05.23295085.

[2]	UN. World Population Prospects 2022, Online Edition. Population by Single Age – Both Sexes 2022. United Nations Department of Economic and Social Affairs. 2022. Accessed at population.un.org/wpp/Download/Standard/Population/.

[3]	Mossong J, Hens N, Jit M, et al. Social contacts and mixing patterns relevant to the spread of infectious diseases. PLoS Med. 2008 Mar 25;5(3):e74.

[4]	IHME. COVID-19 cumulative deaths - Germany. Hans Rosling Center for Population Health, University of Washington, Seattle, US: Institute for Health Metrics and Evaluation; 2021. Accessed at <https://covid19.healthdata.org/germany?view=cumulative-deaths&tab=trend>.

[5]	Keeling MJ, Rohani P. Modeling infectious diseases in humans and animals. Princeton, NJ: Princeton University Press; 2008:159.

[6]	Wölfel R, Corman VM, Guggemos W, et al. Virological assessment of hospitalized patients with COVID-2019. Nature. 2020 May;581(7809):465-469.

[7]	Imai N, Cori A, Dorigatti I, et al. Report 3: Transmissibility of 2019-nCOV London: Imperial College London; 2022. Accessed at <https://doi.org/10.25561/77148>

[8]	Vanni T, Karnon J, Madan J, et al. Calibrating models in economic evaluation: a seven-step approach. Pharmacoeconomics. 2011 Jan;29(1):35-49.
